# Mortality co-benefits of dietary shifts under contrasted trajectories toward anet-zero emission in France by 2050

**DOI:** 10.64898/2026.02.20.26346711

**Authors:** Ines Masurel, Carine Barbier, Christian Couturier, Rémy Slama, Emmanuelle Kesse-Guyot, Kévin Jean

**Affiliations:** Biology department of the Ecole Normale Supérieure (IBENS), CNRS, INSERM, Paris, France; Paris Reasearch in Health, Environment, Climate (PARSEC), Ecole Normale Supérieure (ENS), INSERM, Paris, France; International Centre for Research on the Environment and Development (UMR CIRED), National Center for Scientific Research (CNRS), Montpellier, France; SOLAGRO, Toulouse, France; Nutritional Epidemiology Research Team (EREN), Epidemiology and Statistics Research Center (CRESS), Sorbonne Paris Nord University and University of Paris, Inserm, INRAE, Cnam, Bobigny, France

**Keywords:** Sustainable diets, net-zero emission pathways, health impact assessment, co-benefits, climate change mitigation

## Abstract

**Background:** Food systems—particularly livestock production—account for substantial greenhouse gas (GHG) emissions, while unhealthy diets, characterized by excessive animal-based and insufficient plant-based food consumption, are a major risk factor for all-cause mortality in Europe. Implementing climate mitigation policies related to the GHG emissions of the food system could therefore bring important health co-benefits.

**Methods:** We developed a health impact assessment model based on a life table approach and evaluated the mortality impact of transitions in food consumption through four contrasting scenarios leading to net-zero GHG emissions for France in 2050. We modeled these various dietary shifts and their impacts on premature death by applying the most recent and robust dose-response relationships derived from meta-analyses for 13 food groups.

**Findings:** The different trajectories of dietary shifts translated into a health impact ranging from 19% [uncertainty interval, UI: 17%-21%] to 24% [UI: 21%-26%] of all-cause mortality prevented in 2050 in the French population. Variation in intakes of nuts, red meat, processed meat, whole grains and legumes bring most of the health benefits. Whatever the parameters chosen in the sensitivity analyses, the results remained robust, with about 100,000-200,000 deaths that could be prevented yearly by 2050 in France.

**Interpretation:** The present study highlights the considerable potential health benefits that trajectories toward sustainable diets can bring. These results reinforce the strong convergence of environmental and human health issues in the agri-food sector.

**Funding:** French High Council for the Future of Health Insurance (HCAAM) and the National Agency for Ecological Transition (Ademe).

**Research in context:** *Evidence before this study:* Food systems are a significant contributor to climate change and in parallel, dietary risks are one of the leading causes of all-cause mortality globally, notably in high-income countries such as France. A recent systematic review by Moutet et al. revealed that only two studies evaluating health co-benefits through dietary shifts in net-zero GHG emissions scenarios were published to date. This suggests a convergence and a possible win-win situation between climate change and human health challenges regarding food production and consumption. In order to face the climate crises, governments around the world, and particularly those of the countries historically the largest contributors to climate change, must cut their greenhouse gas emissions to achieve net-zero emission by 2050. Dietary shifts would be a major driver to pursue this objective and could bring important health benefits to the population conducting these changes. For instance, Hamilton et al. showed that dietary changes in line with the Paris Agreements could result in 188 deaths prevented per 100,000 persons in 2040 in Germany and 141 in the UK.

*Added value of this study:* Of the two previously published studies, only one assumed a gradual implementation of changes in diets, combined with a time lag in health effects. We also made these assumptions and considered the gradual change in consumption of thirteen food groups for which recent meta-analyses provided all-cause mortality dose-response relationships with a high level of quality. This study is also among the first to combine nutritional and environmental optimization, through four scenarios; all of which are expected to lead to net-zero emission by 2050 via very contrasting climate change mitigation trajectories. Nevertheless, all of them require a dietary shift toward more plant-based foods. We conducted a health impact assessment for France and showed that achieving net-zero emission by 2050 while considering nutrition references set by national guidelines would provide health co-benefits. Depending on the scenarios, health gains could range from 19% to 24 % of all-cause mortality prevented in the adult French population in 2050, compared to a scenario assuming that we would maintain the current observed dietary habits in the future.

*Implications of all the available evidence:* This study adds to the available evidence that taking action to mitigate climate change is an opportunity to strongly improve public health. Engaging populations in a shift toward a healthier and more sustainable diet could bring major human health and environmental benefits.

## 1 Introduction

The increasing manifestations of climate change require an urgent and systemic transformation of global food systems in order to protect human and ecological health.^1^ Indeed, the agri-food sector is a significant contributor to climate change, representing one quarter to one third of greenhouse gas (GHG) emissions globally.^1,2^ In France, where this sector accounts for 25% of emissions,^3^ animal-based foods contribute to more than half of food-related GHG emissions.^4^ Shifting toward more plant-based diets has been identified by the High-Level Panel of Experts on Food Security and Nutrition as one of the main levers for decarbonizing food systems.^5^

Food systems interconnect with public health, notably through air, soil, and water pollution, as well as food consumption (diet composition and content in harmful substances).^1^ In France, dietary risks are one of the leading causes of mortality, mainly due to diets that are too high in animal-based foods, such as red and processed meat, and too low in plant-based foods, especially whole grains and legumes.^6^ Recent studies show that sustainable diets reduce the risk of chronic diseases, premature deaths, and environmental pressures.^7^ Globally, improved diet quality could help cut food-related GHG emissions by 70% and reduce mortality by 27%, thus providing health co-benefits beyond the mitigation of climate-mediated health risks.^1^

To encourage efforts and identify credible trajectories for reaching net-zero emissions, various governmental and non-governmental organizations have developed scenarios that outline technical and policy solutions for society to achieve that goal. Recent research highlights the significant environmental and health benefits of such trajectories.^8,9^ Nonetheless, very few studies have focused on the share of diet shifts in these scenarios.^10^ There is considerable potential for health gains from trajectories toward carbon neutrality, particularly through changes toward sustainable food consumption. However, different pathways toward net-zero emissions are possible and could thus involve unequal health co-benefits.

We developed a Health Impact Assessment (HIA) model to estimate the health co-benefits on all-cause mortality of different shifts in dietary patterns in the French adult population, in order to contribute to public decision-making on the net-zero carbon transition by 2050 in the agri-food sector.

## 2 Methods

### 2.1 Diets in net-zero emission transition scenarios

Starting from previously defined future diets across four pathways leading to net-zero emissions for France by 2050, we conducted a prospective HIA, following the most recent guidelines for modeling and reporting health effects of climate change mitigation actions.^13^ Each of the transition scenario relied on a different combination of the levers that are available to decarbonize the agri-food sector^11^ (Table 1). The model’s baseline diet was informed by a previous analysis conducted in the French NutriNet-Santé cohort in 2014^14^(Appendix A, Table S1). Then, future diets were developed from a subsample of the French NutriNet-Santé cohort.^15^ Those diets were optimized to minimize the distance to the French nutritional references (considering the daily total energy intake, and a set of macro-nutrients and micro-nutrients) while remaining as close as possible to currently observed diets,^16,17^ and optimized under additional constraints for animal products and imported tropical fruits to lower their GHG emissions^11^(Appendix A, Figure S1).

**Table 1:**
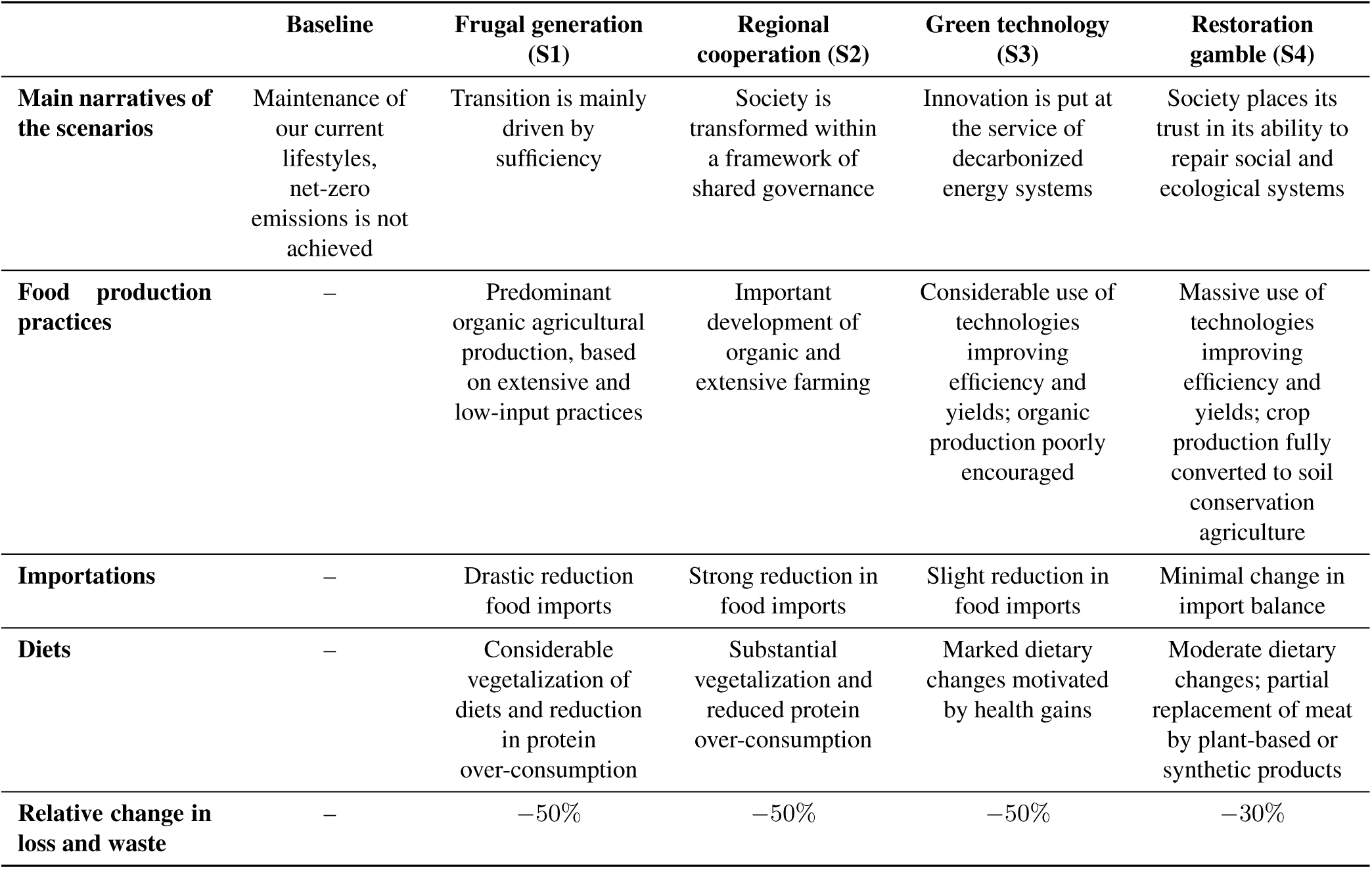
Societal choices and evolutions of the agri-food sector in net-zero emission transition scenarios^11,12^.

We assumed an average diet, homogeneously applied to the population. Intakes (in g/day/person) of animal food products were reduced in all scenarios by 2050. Particularly, reduction in meat (red, processed and white meat) ranges from -31% (S4) to -77% (S1). On the opposite, the proportion of plant-based foods increases in all trajectories, notably through higher consumption of legumes ranging from +68% (S4) to +457% (S1) and nuts ranging from +486% (S4) to +918% (S1) (representing an increase of 10-18 g/day/person). Nevertheless, some food intakes vary non-gradually from S4 to S1, such as whole grains, vegetables and sugar-sweetened beverages. Changes in daily energy intake were not included in the HIA as there is to our knowledge, no meta-analysis reporting relationships between energy intake and all-cause mortality. Moreover, the total energy intake varies little between the baseline and the scenarios, and between the different scenarios themselves (Table 2, Table S3, Figure S2, Figure S3).

**Table 2:** Food intakes (g/day/person) and total energy intake (kcal/day/person) in the current and 2050 net-zero emission scenarios diets and relative changes from the current diet^11^.

|  | Current diet | S1 |  | S2 |  | S3 |  | S4 |  |
| --- | --- | --- | --- | --- | --- | --- | --- | --- | --- |
| Food group | Intake | Intake | Relative shift from current diet | Intake | Relative shift from current diet | Intake | Relative shift from current diet | Intake | Relative shift from current diet |
| Red meat | 98 | 21 | −79% | 34 | −65% | 48 | −51% | 61 | −38% |
| Processed meat | 37 | 3 | −91% | 9 | −77% | 11 | −69% | 24 | −34% |
| White meat | 26 | 13 | −48% | 19 | −26% | 26 | 3% | 26 | 2% |
| Dairy | 269 | 107 | −60% | 142 | −47% | 164 | −39% | 219 | −19% |
| Fish | 32 | 19 | −42% | 24 | −17% | 31 | −4% | 41 | 28% |
| Eggs | 11 | 32 | 189% | 27 | 143% | 23 | 108% | 15 | 39% |
| Fruits | 244 | 136 | −44% | 155 | −37% | 172 | −29% | 241 | −1% |
| Nuts | 2 | 20 | 918% | 17 | 736% | 13 | 533% | 12 | 486% |
| Vegetables | 280 | 380 | 36% | 360 | 29% | 337 | 20% | 347 | 24% |
| Legumes | 15 | 81 | 457% | 58 | 299% | 46 | 212% | 25 | 68% |
| Whole grains | 11 | 33 | 199% | 30 | 168% | 27 | 145% | 43 | 284% |
| Refined grains | 178 | 154 | −14% | 156 | −13% | 151 | −15% | 142 | −20% |
| Sugar-sweetened beverages | 174 | 126 | −28% | 132 | −24% | 142 | −18% | 136 | −22% |
| **Total energy intake (kcal)** | 2005 | 1780 | −11% | 1758 | −12% | 1719 | −14% | 1870 | −7% |

### 2.2 All-cause mortality risks

In line with previous studies, thirteen food categories were selected (red meat, processed meat, white meat, fish, eggs, dairy, fruits, vegetables, legumes, nuts, whole grains, refined grains and sugar-sweetened beverages), for which recent meta-analyses provided dose-response relationships for all-cause mortality with a high level of evidence on the AMSTAR-2 scale^18–27^ (Figure 1). The quality of evidence for each food group was previously rated using the NutriGrade scoring system^28^ (Table S4). The relative risk (RR) of a diet, defined as a specific intake of each of the thirteen food groups, was calculated as the product of the RRs associated with each food group (Appendix E).

**Figure 1:**
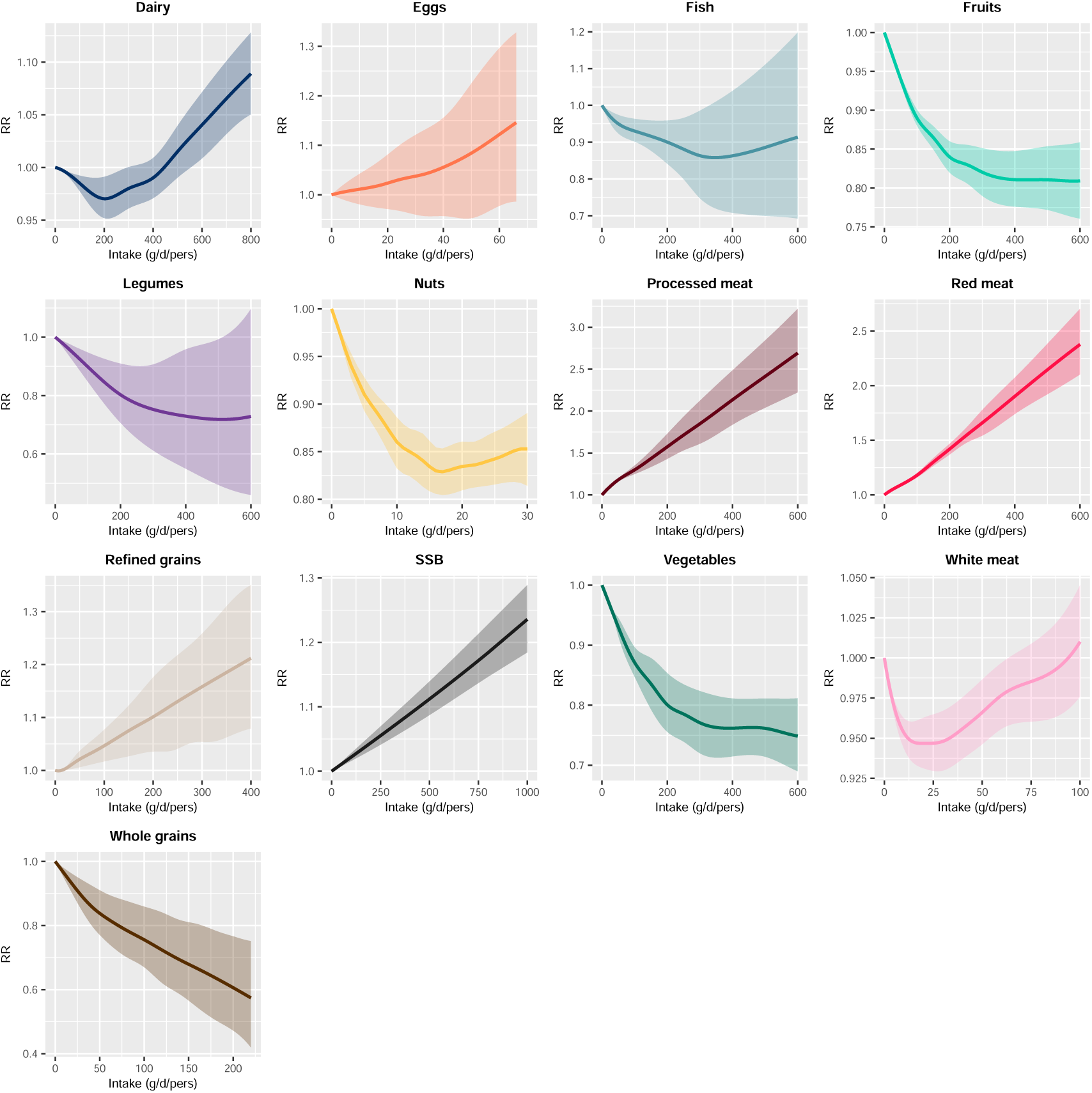
All-cause mortality dose-response relationships of each food group included in the DOUGLAS model. RR = Relative Risk, SSB = Sugar-Sweetened Beverages

The studies selected into the meta-analyses adjusted their analyses for confounding factors, including intake of other food groups. However, due to differences in food categories included in adjustments, overlapping benefits of different food groups may occur, thus overestimating the effects of each food group. In line with previous publications, we accounted for a 25% attenuation in risk estimates of each food group on mortality^18,19^ (Appendix D).

### 2.3 Temporality of the health impacts

The timing of health impacts is influenced by the implementation dynamics of diets and the time lag between a change in diet and its health impacts. Shifts from current to modeled diets were assumed to follow a S-curve from 2025 to 2050 (Appendix B). All-cause mortality associated with diet is almost exclusively related to non-communicable chronic diseases (mainly cardiovascular disease, type II diabetes and cancer).^29^ Therefore, in order to take the development period of these mortality related diseases into account, a linear 10-year time to full effect was used in the model, as previously described.^18^ Thus, the RR associated with each food group in a given year was calculated as a weighted mean of year-specific RRs over the previous 10 years, with weights increasing linearly from the most recent to the oldest year, consistent with the assumed time lag (Appendix E).

### 2.4 Health impact model

French adult population size and mortality projections were obtained from publicly available sources.^30^ Changes in mortality due to dietary shifts were calculated among the adult (≥ 18y) population only. The number of deaths prevented by the diet shift of a transition scenario was calculated as follows:

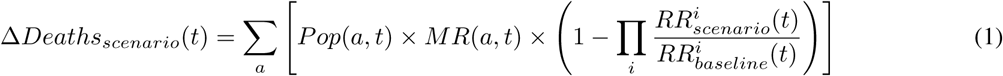

Where *a* is the age, *t* is the year, and *i* is the food group. *Pop* denotes the population size and MR the mortality rate.

Based on the yearly projected life expectancy, we also calculated the life expectancy gain and the years of life lost (YLL) averted by shifting diet (Appendix E). For each scenario, we also computed the contribution of each food group to the total prevented mortality, as the percentage of mortality attributable to variation in consumption of each of them (Appendix F).

### 2.5 Uncertainty and sensitivity analyses

Uncertainty around the estimated effects of dietary changes on mortality was quantified using a Monte Carlo simulation approach. For each food group, RRs and their associated 95% confidence intervals were obtained from published meta-analyses and used to define a log-normal probability distributions, from which we randomly sampled values that were used to estimate the model outcomes. A total of 1,000 combinations of RR values were used, and the resulting uncertainty intervals (UI) were defined by the 2.5*^th^* and 97.5*^th^* percentiles of the simulated outcomes.

The main analysis assumed a cosine interpolation implementation of diets from 2025 to 2050, as well as a 10-year linear time to full effect and a 25% reduction in mortality risk for each food group. Different sensitivity analyses were performed. The first one explored a transition in diet starting in 2016, right after the Paris Agreement; the second one explored an immediate dietary implementation in 2025. Then, we conducted analyses with no attenuation or a 50% attenuation in the RRs. Lastly, we used either an immediate or a 20-year long time to full effect.

## 3 Results

### 3.1 Quantitative health impact assessment

As compared to maintaining the current diet, all transition scenarios result in an overall improvement of health (Figure 2, Table 3). In 2040, 15 years after the initiation of the diet change, all-cause mortality in the French adult population would be decreased by 7.2% [UI: 6.4%-7.9%] in Scenario 3 (S3) and up to 10.2% [Uncertainty Interval, UI: 9.1%-11.4%] in S1. Impacts were larger in 2050, yearly all-cause mortality would be decreased from 19.2% [UI: 17.3%-21.2%] in S3 to 24.2% [UI: 21.6%-26.9%] in S1, corresponding to a number of yearly prevented deaths ranging from 152,000 [UI: 137,000-168,000] in S3 to 191,100 [UI: 171,000-213,000] in S1. In all scenarios, the dietary targets assumed by the prospective exercise are met by 2050. Given the time lag between food intakes and health effects, the benefits provided by the diet changes continued to progress after 2050, reaching a plateau of 26.9% [UI: 24.0%-30.1%] of yearly all-cause mortality prevented in S1 in 2060.

**Figure 2:**
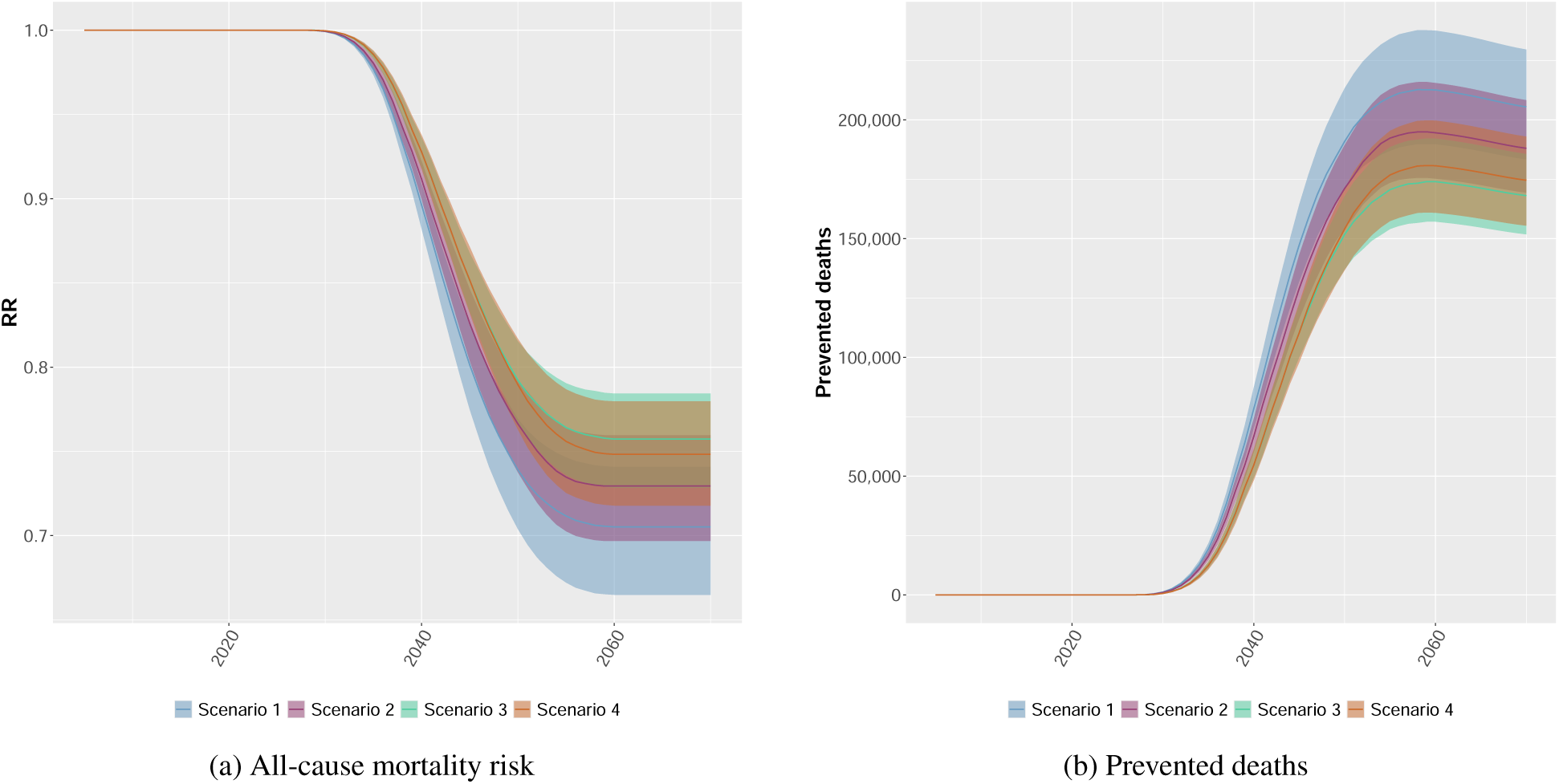
Time variation in (a) the Relative Risk (RR) associated with all-cause mortality and (b) the number of deaths prevented yearly through diet shifts in transition scenarios in the adult French population compared to keeping the current diet.

**Table 3:** Heath co-benefits of diet shifts in different transition scenarios.

| Scenario | 2040 | 2050 | 2060 |
| --- | --- | --- | --- |
| *Deaths prevented (UI, in thousands)* |  |  |  |
| S1 | 78.3 (70.3-87.3) | 191 (171-213) | 213 (190; 238) |
| S2 | 66.7 (60.1-74.1) | 171 (154-189) | 195 (175-216) |
| S3 | 54.6 (48.2-61.2) | 152 (137-168) | 174 (157-192) |
| S4 | 55.0 (48.0-61.0) | 154 (137-171) | 181 (161-200) |
| *Years of life saved (UI, in thousands)* |  |  |  |
| S1 | 855 (763-957) | 2,100 (1,850-2,370) | 2,250 (1,970-2,560) |
| S2 | 724 (650-807) | 1,850 (1,660-2,070) | 2,030 (1,810-2,280) |
| S3 | 589 (530-655) | 1,620 (1,450-1,810) | 1,790 (1,600-2,000) |
| S4 | 589 (518-662) | 1,650 (1,450-1,850) | 1,870 (1,650-2,090) |
| *Life expectancy gained (UI, in months)* |  |  |  |
| S1 | 8.96 (7.99-10.0) | 22.8 (20.1-25.9) | 25.3 (22.2-28.8) |
| S2 | 7.58 (6.80-8.45) | 20.1 (18.0-22.5) | 22.9 (20.3-25.7) |
| S3 | 6.16 (5.54-6.86) | 17.6 (15.7-19.7) | 20.2 (18.0-22.6) |
| S4 | 6.16 (5.41-6.93) | 17.9 (15.7-20.1) | 21.0 (18.5-23.6) |

### 3.2 Food group contributions

The decrease in mortality was primarily due to an increase in the consumption of nuts (mortality decreased by -7.3% [UI: -6.0%- -8.7%] in S4 to -9.7% [UI: -8.3%- -11.1%] in S1), whole grains (from -3.7% [UI: -1.9%- -5.3%] in S3 to -6.8% [UI: -3.5%- -9.7%] in S4) and legumes (from -0.6% [UI: -0.3%- -0.9%] in S4 to -4.1% [UI: -2.1%- -6.2%] in S1) and a decrease in consumption of red meat (from -3.4% [UI: -3.1%–3.7%] in S4 to -6.3% [UI: -6.1%–7.4%] in S1) and processed meat (from -2.2% [UI: -2.0%–2.3%] in S4 to -6.4% [UI: -5.8%–6.9%] in S1). Conversely, we observed an elevation in mortality due to reduced consumption of fruits (mortality increased by 0.02% [UI: 0.01%-0.04%] in S4 to 2.6% [UI: 1.7%-3.2%] in S1) and fish (from 0.07% [UI: 0.04%-0.10%] in S3 to 0.81% [UI: 0.42%-1.2%] in S1), and an increased consumption of eggs (from 0.25% [UI: -0.30%-0.88%] in S4 to 1.6% [UI: -1.3%-4.7%] in S1). Overall, these slight negative impacts on mortality were largely offset by the benefits brought by changes in intake of other food groups in future diets (Figure 3).

**Figure 3:**
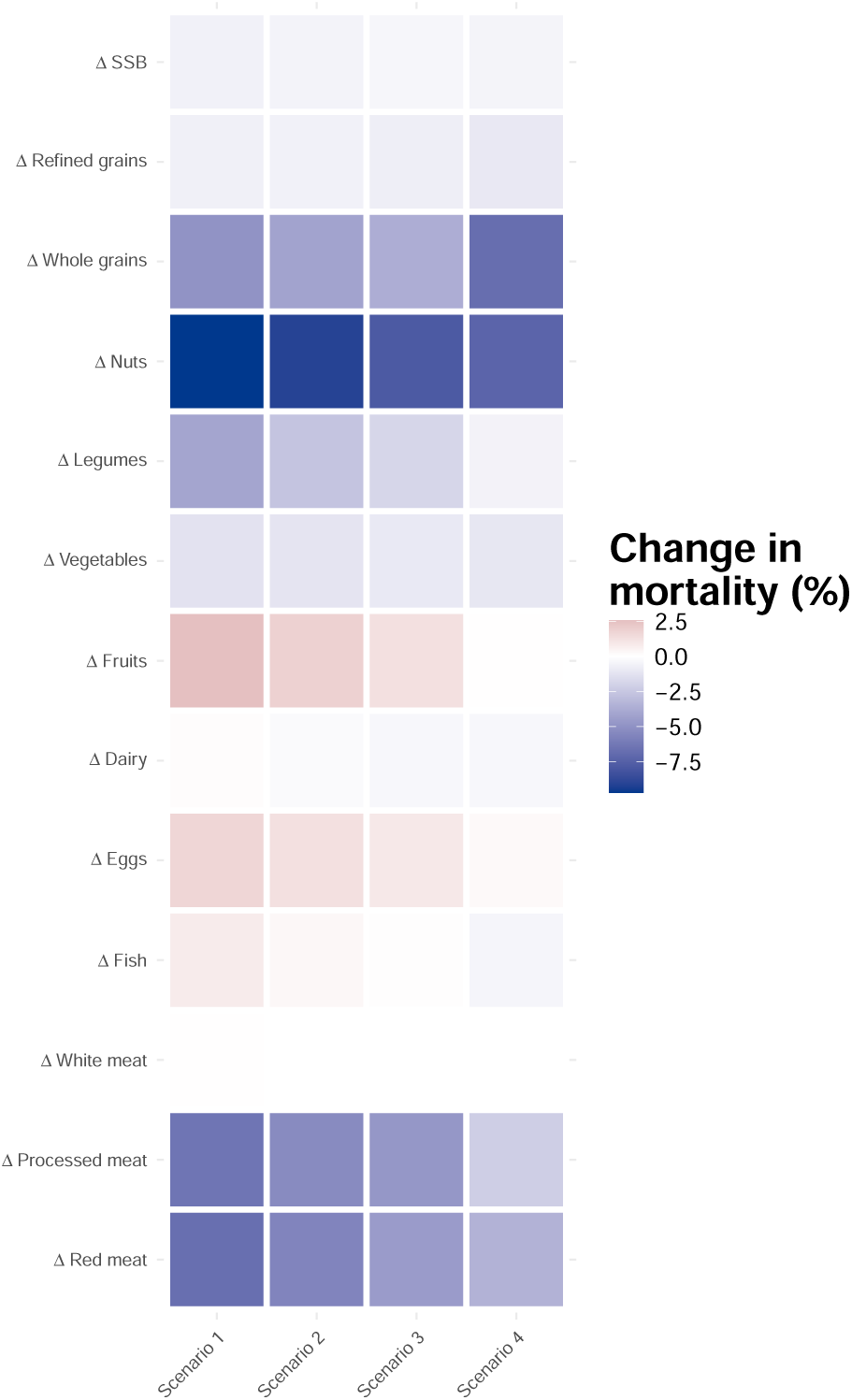
Change in 2050 yearly all-cause mortality attributed to the variation in consumption of different food groups within transition scenarios compared to maintaining the current observed diet in France. SSB: Sugar-sweetened beverages.

### 3.3 Sensitivity analyses

An early transition would lead to more immediate health improvements, but it wouldn’t change the ranking of the scenarios. If such a transition had been implemented in 2016, right after the Paris Agreement, between 6,000 and 10,000 deaths could have been avoided in 2026, depending on the scenario. As expected, assuming an immediate implementation of the dietary changes in 2025 would result in higher numbers of prevented deaths (relative burden deviation from main analysis in S1, 2040: +167.8% [UI: 166.7-168.5] %; 2050: +12.5% [UI: 12.2-12.5] %). Then, assuming an immediate effect on mortality of any intake would considerably increase the number of prevented deaths (S1 in 2040: +112.0% [UI: 111.0-113.4] %; 2050: +12.6% [UI: 12.3-12.6] %). In addition, the maximum decrease in all-cause mortality risks would be reached as soon as the dietary shift ends, in 2050. Conversely, assuming a 20-year period for the time to full effect would delay the health outcomes (S1 in 2040: -70.0% [UI: -70.1- -69.8] %; 2050: -37.0% [UI: -37.1- -37.0] %), with the maximum decrease in all-cause mortality risks reached in 2070. Considering no confounding effects between food intakes, would increase the estimates (S1 in 2040: +33.9% [UI: 34.3-33.8] %; 2050: +31.1% [UI: 30.1-31.7] %). On the contrary, considering a more important confounding effect between food groups and attenuating their risk effect by 50% would decrease the health outcomes of the same order of magnitude. Finally, an analysis with no change in demographic structure showed moderately lower results (Figure 4, Figure S4, Table S5, Table S6). An additional analysis estimated that the health gains obtained through the shift towards S1 diet represented about half of those that could be expected from a nutritionally optimized diet (Appendix H).

**Figure 4:**
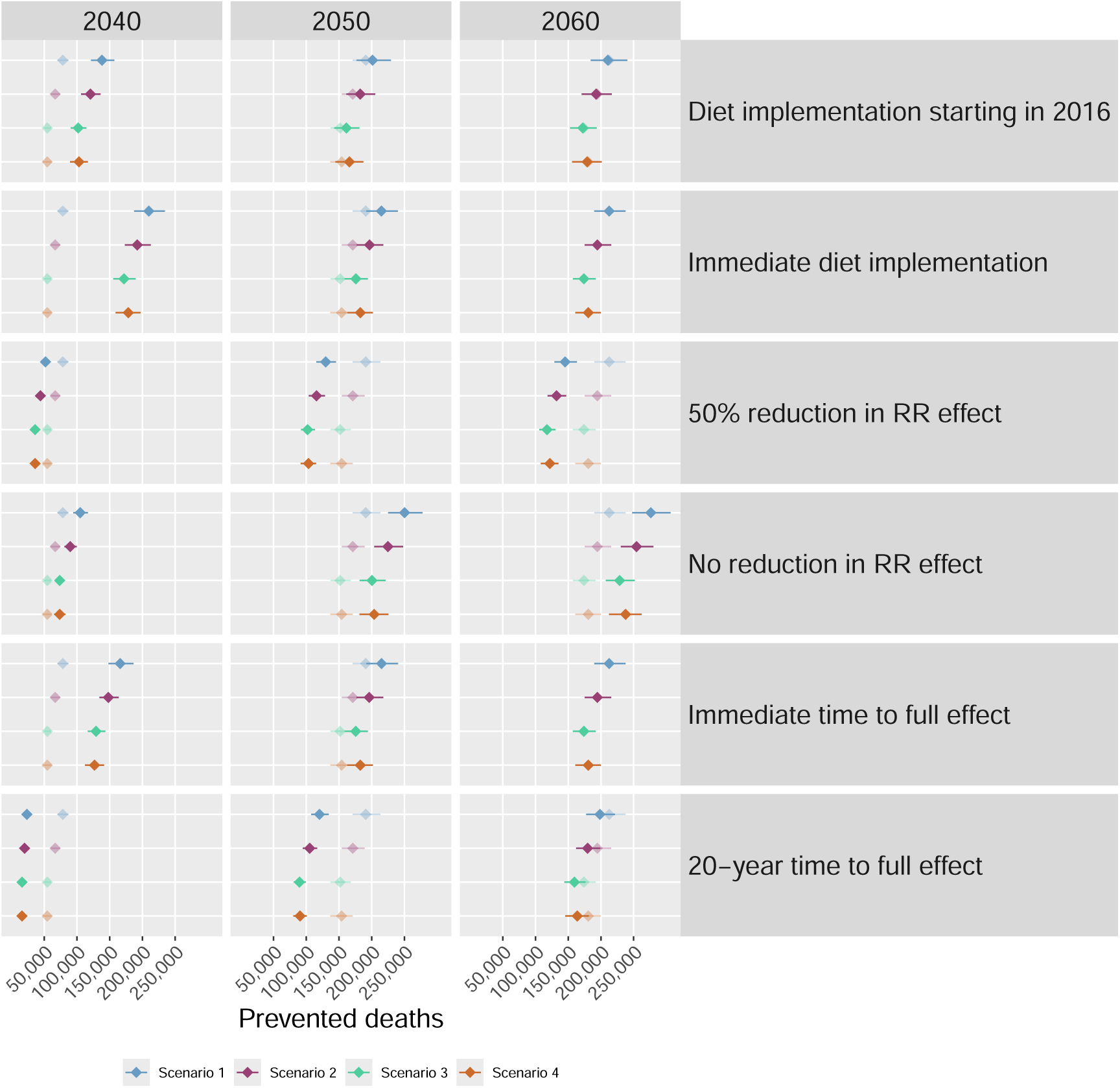
Number of deaths prevented in 2040, 2050 and 2060 under different assumptions. The points in transparency represent the main analysis results.

## 4 Discussion

In each of the climate change attenuation scenarios assessed, dietary changes consistent with net-zero emissions imply a dramatic reduction in all-cause mortality, ranging from 152,000 [UI: 137,000-168,000] to 191,000 [UI: 171,000-213,000] deaths prevented yearly by 2050 compared to maintaining the current diet, corresponding to 19.2% [UI: 17.3%-21.2%] to 24.2% [UI: 21.6%-26.9%] of all-cause mortality prevented in 2050. Whatever the parameters chosen in our sensitivity analyses, the results remained robust, with about 100,000 to 200,000 deaths that could be prevented by 2050 in France. All scenarios imply dietary shifts towards more plant-based foods with contrasted magnitude. In particular, nuts intake is increased by 486% to 918% and meat intake is reduced by -31% to -77% compared to the current diet. Dietary transitions that involve the greatest shift from animal foods to plant-based foods, first S1 then S2, appear to be the most beneficial for health, in line with the literature.^1^

The highest intake of animal products is found in S4, particularly red meat and processed meat, which considerably increases the risk of all-cause mortality.^25^ However, the health benefits estimated in S3 and S4 are very close, with the health benefits of S4 rising above those of S3 shortly before 2050. Indeed, the number of life years saved in 2050 reaches 1,650,000 [UI: 1,450,000-1,850,000] in S4 and 1,620,000 [UI: 1,450,000-1,810,000] in S3, corresponding to a gain in life expectancy of 17.9 [UI: 15.7-20.1] and 17.6 [UI: 15.7-20.1] months, respectively. This can be explained by the process of environmental and nutritional optimization of diets, which places fewer constraints on S4 than on S3, meaning that the diet in S3 is less favorable than in the other scenarios for certain food groups, particularly vegetables, whole grains, and sugar-sweetened beverages.^11^

Previous studies performed HIA of changes toward diets meeting nutritional constraints, like Eustachio et al., who showed that meeting the recommendations on fruit and vegetable intake in the UK would allow a gain in life expectancy at birth of 7-8 months.^31^ Other recent studies carried out HIA applied to low GHG emissions diet shifts, such as Milner et al., who evaluated the health outcomes of a shift toward an average diet in the UK complying with the World Health Organization (WHO) nutritional targets while lowering its GHG emissions by 17%, resulting in an average life expectancy gain of 8 months.^32^ In comparison, the transition scenarios evaluated here all target net-zero emissions by 2050 and thus imply a larger cut in GHG emissions,^11^ leading to more significant changes in diets and explaining that the results presented here are higher in magnitude. A recent observational study conducted in a French cohort showed that higher adherence to some dietary guidelines accounting for sustainability led to more than 35,000 averted deaths a year.^33^ Nevertheless, these environmental considerations are far from sufficient and must include limits on total meat consumption.^34^ Two other recent publications performed a multi-sectorial HIA - including dietary changes

- of net-zero emission pathways, resulting in 489,000 life years preserved from 2021 to 2050 in England and Wales^9^ and 143,000 deaths prevented in 2040 in Germany and 100,000 in the UK.^8^ Overall, our results appear to be of the same magnitude as the available evidence. The large potential health benefits we documented here are also consistent with the fact that dietary risks are estimated to be the second factor of all-cause mortality in France.^6^ Fadnes et al. recently showed that shifting from a typical Western diet to a longevity-optimized diet would bring a life expectancy gain of about 8 and 9 years for 20-year-old French females and males, respectively.^19^ The evaluated benefits are much greater than those estimated here, as they stem from the immediate adoption of diets that significantly reduce or even eliminate high-risk food groups, such as red and processed meats.

When conducting nutritional HIA, accounting for possible residual correlations between food groups is essential to avoid over or underestimating the health impacts.^35^ However, there is little literature on the exact magnitude of attenuation to be applied to account for this effect. Previous studies carried out their projections with a 25% reduction,^18,19^ but other recent publications didn’t apply any attenuation factor.^8,9,31^ In addition, it is likely that correlations differ between the food groups, but we assumed the same attenuation factor. However, it is also possible that the effects of each food group have been over-adjusted in meta-analyses. Here, we applied a conservative 25% attenuation factor in each RR.

In our projections, the temporality of health co-benefits is determined by the dynamics of the implementation of diets and the time lag between food consumption and it’s impacts on health. We assumed a gradual implementation of diets (Appendix B, Figure S2). Changes toward more sustainable diets in a population rely on several behavioral mechanisms and socio-economic factors and thus can be modeled as a logistic function.^36^ Using such diet implementation dynamics, assuming social influence, could represent a further improvement to our model. Mortality related to food consumption is almost exclusively due to non-communicable diseases like cancers, cardiovascular diseases, and type II diabetes. Evidence from other fields, especially from tobacco consumption^37^ and air pollution^38^ epidemiology, has described the latency in these diseases’ natural history. Currently, the time lag between changes in diets and effects on mortality risk attributable to these diseases is not accurately documented. Few studies based on empirical evidence describe lags reaching a maximum impact after 10 years for cardiovascular diseases and type II diabetes^39^ but some other studies show important changes in cardiovascular diseases after only 5 years of intensive dietary changes^40^ or even less.^41,42^ The time lags between dietary changes and the subsequent effects on mortality associated with cancer are likely to be much longer - probably several decades, with little change in the first 10 years.^32,43–45^ Cause-specific mortality time lag functions have been used in other recent studies^8,9,31,32^ and informed as much as possible by empirical evidence in nutritional or smoking epidemiology.^39,41,44–46^ This calls for more observational studies focused on the temporality of dietary shifts on health and on the correlations between food intakes to better inform our model. Combined, the gradual implementation of diets and the time to full effect imply that the first health benefits to be observed 5 years after the beginning of the diet shift at the earliest. Highlighting that such latency in health benefits is expected is crucial for informing public policy decisions. However, this should not prevent governments from committing to transitioning toward net-zero carbon emissions. Our sensitivity analyzes reveal that if such a dietary transition had been initiated as soon as 2016 (i.e., after the Paris Agreements), up to 10,000 deaths could have been prevented as of 2026, emphasizing the importance of the early implementation of climate policies.

Other climate policies have been assessed for their health impacts. A recent study using ADEME scenarios for transportation, found that increasing walking and cycling could lead to substantial health gains (up to 25 000 deaths prevented by 2050 for the most active scenario), whereas a scenario focused on replacing combustion cars with electric ones but reducing walking would yield worse health outcomes.^47^ Recent multi-sectorial net-zero emission trajectory studies show that health benefits differ widely between sectors: In Germany in 2040, the number of deaths prevented due to reduction in air pollution would be about 15,600 those due to increase in active travel would be about 5,600 and 143,000 would be attributed to changes toward more sustainable diets.^8^ Over the 2021-2050 period in England and Wales, 891,000, 287,000 and 489,000 years of life could be gained, in the same sectors respectively.^9^ Therefore, impacts of dietary shifts may constitute one of the highest potentials for health co-benefits of climate policies. But integrating all these levers into ambitious climate policies could bring even greater health benefits.

Our study has several limitations. First, we assumed that an average diet would be applied homogeneously to the population. These average diets reflect a distribution of different consumer profiles (Table S2). Yet, as we did not take demographic characteristics into account in our model, these different diets are distributed in the same way across ages, genders and socioeconomic categories. This lack of heterogeneity, which is common in HIA studies, could affect our estimates, though it is not clear at this time to which extent. Future work will consider different dietary patterns across socioeconomic profiles. Then, the quality of evidence of the relationships between intake and all-cause mortality was rated as “*very low*” by the NutriGrade scoring system for some food groups included in the model, such as eggs and white meat, but these are not the food groups contributing the most to the results. The present study was based on the most recent meta-analysis for which the quality was ranked as “*high*” on the AMSTAR-2 grading scale.^26^ Additionally, although food systems are linked to multiple environmental pressures, only GHG emissions have been reduced here. Moreover, a 2014 diet derived from the French NutriNet-Santé cohort study was taken as the baseline of our model, but we set the beginning of the HIA in 2025. A recent publication highlighted changes in the dietary habits of the French population over the past decade,^48^ but those changes remained limited compared to those expected under the climate change mitigation scenarios investigated here. This study also presents several strengths. Although two previous studies performed dietary HIA in net-zero emission scenarios, only one of them accounted for both a gradual implementation of diets and a time lag in their health effects. We also made these assumptions and considered the gradual change in consumption of thirteen food groups. This study is also among the first to combine nutritional and environmental optimization throughout four scenarios, all of which lead to net-zero emission by 2050 via very contrasting climate change mitigation trajectories.

Approaching dietary transition from the perspective of climate change may be simplistic; other environmental issues are linked to agri-food systems.^1^ Potential conflicts may arise between environmental factors, in particular between GHG emissions and water use in certain food productions.^49^ Other conflicts over nutrition may also emerge. It is important to clarify which healthy and sustainable plant-based products should be substituted for animal-based products, especially since plant-based foods include both healthy and unhealthy foods, such as ultra-processed foods and/or fatty and sweet products.^50^ In addition, a higher proportion of plant-based food might increase the pesticide residues exposure.^51^ Nevertheless, diets with a low overall environmental impact were assessed to provide important health benefits.^7^

## 5 Conclusion

Achieving net-zero emissions imply a substantial shift toward more plant-based diets. Promoting such dietary transitions among the French population by 2050—aligned with both environmental and public health goals—could yield significant health co-benefits alongside environmental gains. The results presented here reflect the combined pursuit of these objectives and offer valuable insights for guiding public policy and identifying optimal transition strategies. They emphasize the significant opportunity to improve population health and advance efforts toward net-zero emission, reinforcing the urgency of implementing integrated climate and health policies. Any further delay would not only increase climate-related risks but also result in missed health gains.

## Data Availability

All data produced in the present study are available upon reasonable request to the authors

## Contributors

KJ, EKG, and IM contributed to the conceptualization of the study, providing the overarching ideas and research goals. KJ and EKG supervised the analyses conducted by IM. IM designed the model, wrote the code, conducted all analyses, and prepared the initial draft of the manuscript. CB and CC provided the dietary data of the 2050 scenarios used in the study. All authors participated in reviewing and editing the manuscript, critically revised the article for content, and approved the final version for publication.

## Data sharing

Data and R scripts used in this study are available here: https://github.com/Inesmsrl/DOUGLAS

## Appendix

### A Current and future diets

In 2022, the results of a prospective exercise exploring trajectories to achieve carbon neutrality by 2050 in the agri-food sector in metropolitan France - including shifts in diets - were described in the SISAE report.^11^ Four scenarios were drawn up, involving highly contrasted societal changes in terms of socio-economic systems, governance, use of technologies and lifestyles. The “Frugal generation” scenario (S1) essentially relies on demand reduction through individual behavior changes, the “Regional cooperation” scenario (S2) is based on changes in social organization practices, promoting sharing and exchanges between the different public and private societal stakeholders, the “Green technologies” scenario (S3) mainly addresses climate challenges by scaling up existing technologies and the “Restora-tion gamble” scenario (S4) consists in managing, or even repairing, the impacts of our lifestyles on social and ecological systems, by relying on certain technologies that are not yet mature.^12^ This last scenario is considered the business-as-usual scenario.^11^ Each of the transition scenario uses a different combination of the levers that are available to decarbonize the agri-food sector.^11^ The French Agency for Ecological Transition (ADEME) has then included these results on the agri-food sector in a report describing narratives aimed at achieving net-zero carbon by 2050 in metropolitan France for all sectors combined. For the present study, we used an additional scenario that was used as the baseline, in which we consider the current lifestyle across years.^14^

This document describes the current diet chosen as the baseline of our study and relates how the diets in the 2050 net-zero emission scenarios were defined as part as the SISAE project.^11^

#### A.1 Current observed diet

The current diet in the French population was defined from a NutriNet-Santé cohort study.^14^ This study included 28,245 participants who answered an organic food frequency questionnaire (Org-FFQ) in 2014. Five groups (quintiles) were defined according to the level of organic consumption. The socio-demographic characteristics of the Q1 group were the closest to the French population. Thus, Q1 food intakes were chosen as the current diet in the French population.

**Table S1:**
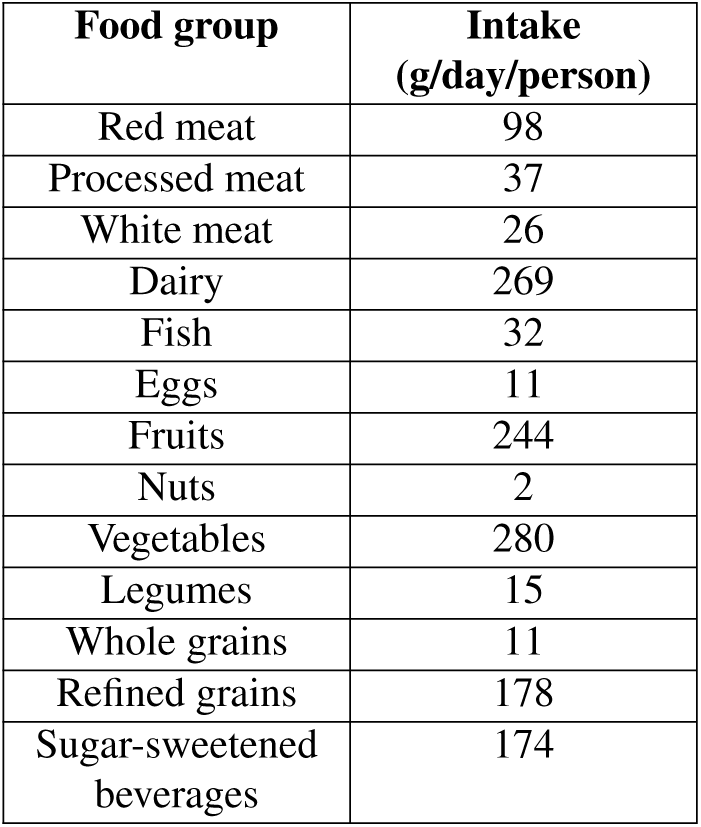
Intakes of the different food groups included in the health impact assessment model, in the current observed diet in the French population (2014)

### A.2 Future diets

#### A.2.1 Observed diets

The 2050 dietary targets were developed from a subsample of the French NutriNet-Santé cohort. From June to December 2014, a semi-quantitative food frequency questionnaire (FFQ-bio) to quantify dietary intakes, and particularly intakes of organically produced foods over the past 12 months, was completed by about 35,000 participants. Six dietary profiles were identified: little (meat-3), moderate (meat-2) and large (meat-1) meat consumers, pescetarians, vegetarians and vegans^15^ (Table S2).

#### A.2.2 Optimization under nutritional and environmental constraints

As previously described,^16^ non-linear optimization models were implemented to define, for each profile, a daily diet in line with the French nutritional references^17^ for men, women under 55 years old and women over 55 years old, as the nutritional recommendations differ according to these groups. In addition, the model included acceptability constraints for food items and 37 food groups (high threshold set at the 95th percentile of group distribution). The nutritional optimization model operated on a food-by-food basis, mobilizing the most relevant foods within groups to meet the constraints imposed, while minimizing the distance from observed consumption.^15^ Thus, the model could select foods with a higher nutritional density, such as fruit rather than fruit juice. Indeed, juices are higher in sugar and lower in fiber, so the model would eliminate them in favor of fruit. This helped to reduce the daily volumes consumed without impacting on the nutritional intake.

The nutritionally adequate diet intakes (in g/day/person) were matched with the statistical values of food availability provided by the Food and Agriculture Organization of the United Nations (FAO). A conversion matrix was used to convert diets described in BioNutriNet format with 264 foods (white bread, sweet crepes…), to the commodity nomenclature, based on about a hundred food or agricultural commodities (wheat, eggs, milk, sugar…). To do so, a table of the ingredients that make up each food (flour, cheese, etc.) was used to obtain the commodities. Coefficients accounting for economic and biophysical allocation as well as the loss or gain of mass (cooking, peelings, etc.) were also considered. For each commodity, the quantities consumed per year at national level were obtained, allowing a comparison with the quantities delivered to the market.

2050 diets were defined in a second phase. First, future agricultural systems were modeled with the MoSUT tool (Modèle systémique d’utilisation des terres) developed by SOLAGRO. MoSUT can be used to adjust food supply and demand, taking into account the evolution of different production systems and requirements for food and non-food products derived from agricultural and forestry biomass. It can also take into account assumptions about international trade. MoSUT is a biophysical back casting model, which identifies one or more possible trajectories for achieving a set of predefined objectives. The exercise is iterative, with adjustments to assumptions and results and bottom-up. Future production systems were imagined on the basis of current representative systems projected to 2050 through the integration of agronomic practices and technical evolutions clearly identified and documented in scientific and technical literature. Assumptions about land use, agricultural production, international trade, greenhouse gas emissions and bio-energy use were used to model 2050 agricultural systems, resulting in food commodity consumption data at the national level for each scenario that were used in the environmental optimization of the previously defined diets. Two additional constraints were considered in order to limit greenhouse gas emissions during farming and transport: a reduction in the consumption of animal products, with a priority given to ruminant meat and a reduction in the consumption of imported tropical fruits, along with a reduction in the consumption of citrus juices.

#### A.2.3 2050 scenarios - population average diets

6 optimized diets were defined with the following weights given to each population group: 50% men, 27% women under 55 years old and 23% women over 55 years old. In each scenario, different adoption rates of these diets were assumed. The distribution (in %) of observed and optimized diets in the different scenarios is summarized in Table S2. As a result, the average diets in the 2050 net-zero emission scenarios are described in Table S3.

**Table S2:** Distribution (%) of observed and optimized diets across different scenarios.

|  | **Current diet** | **Frugal**<br>**generation**<br>**(S1)** | **Territorial**<br>**cooperation**<br>**(S2)** | **Green**<br>**technologies**<br>**(S3)** | **Restauration**<br>**gamble (S4)** |
| --- | --- | --- | --- | --- | --- |
| Meat-1 | 57 | 6 | 15 | 20 | 43 |
| Meat-2 | 29 | 3 | 7 | 10 | 22 |
| Meat-3 | 10 | 1 | 3 | 4 | 8 |
| Pescetarian | 1 | 0 | 1 | 0 | 1 |
| Vegetarian | 2 | 0 | 0 | 0 | 2 |
| Vegan | 1 | 0 | 0 | 0 | 1 |
| Meat-1 optim | 0 | 11 | 14 | 33 | 14 |
| Meat-2 optim | 0 | 17 | 24 | 37 | 10 |
| Meat-3 optim | 0 | 33 | 25 | 15 | 0 |
| Pescetarian optim | 0 | 1 | 1 | 0 | 0 |
| Vegetarian optim | 0 | 22 | 9 | 5 | 1 |
| Vegan optim | 0 | 6 | 1 | 1 | 0 |

**Table S3:**
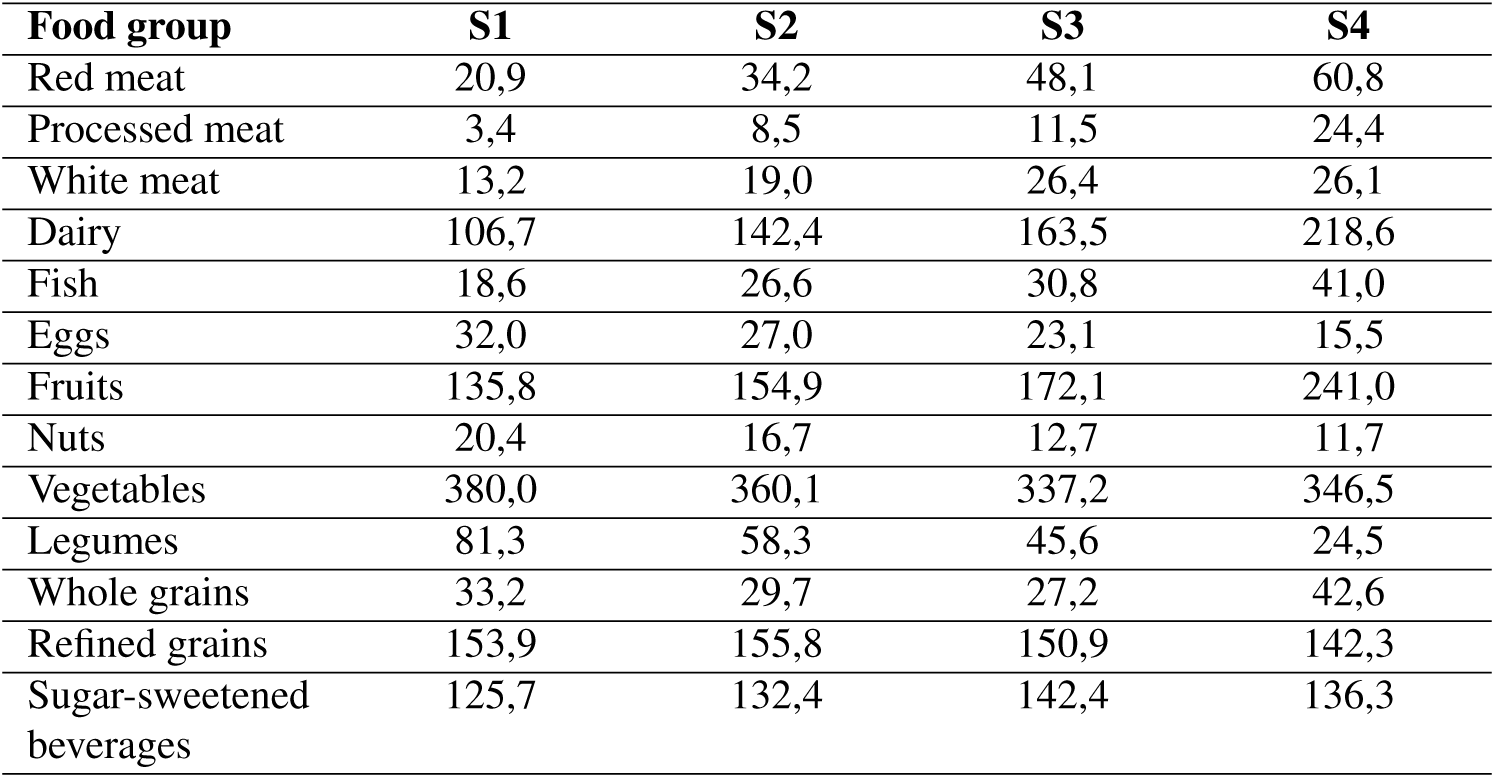
Intakes (g/day/person) of the different food groups included in the health impact assessment model, in each 2050 transition scenario.

| **Food group** | **S1** | **S2** | **S3** | **S4** |
| --- | --- | --- | --- | --- |
| Red meat | 20,9 | 34,2 | 48,1 | 60,8 |
| Processed meat | 3,4 | 8,5 | 11,5 | 24,4 |
| White meat | 13,2 | 19,0 | 26,4 | 26,1 |
| Dairy | 106,7 | 142,4 | 163,5 | 218,6 |
| Fish | 18,6 | 26,6 | 30,8 | 41,0 |
| Eggs | 32,0 | 27,0 | 23,1 | 15,5 |
| Fruits | 135,8 | 154,9 | 172,1 | 241,0 |
| Nuts | 20,4 | 16,7 | 12,7 | 11,7 |
| Vegetables | 380,0 | 360,1 | 337,2 | 346,5 |
| Legumes | 81,3 | 58,3 | 45,6 | 24,5 |
| Whole grains | 33,2 | 29,7 | 27,2 | 42,6 |
| Refined grains | 153,9 | 155,8 | 150,9 | 142,3 |
| Sugar-sweetened<br>beverages | 125,7 | 132,4 | 142,4 | 136,3 |

**Figure S1:**
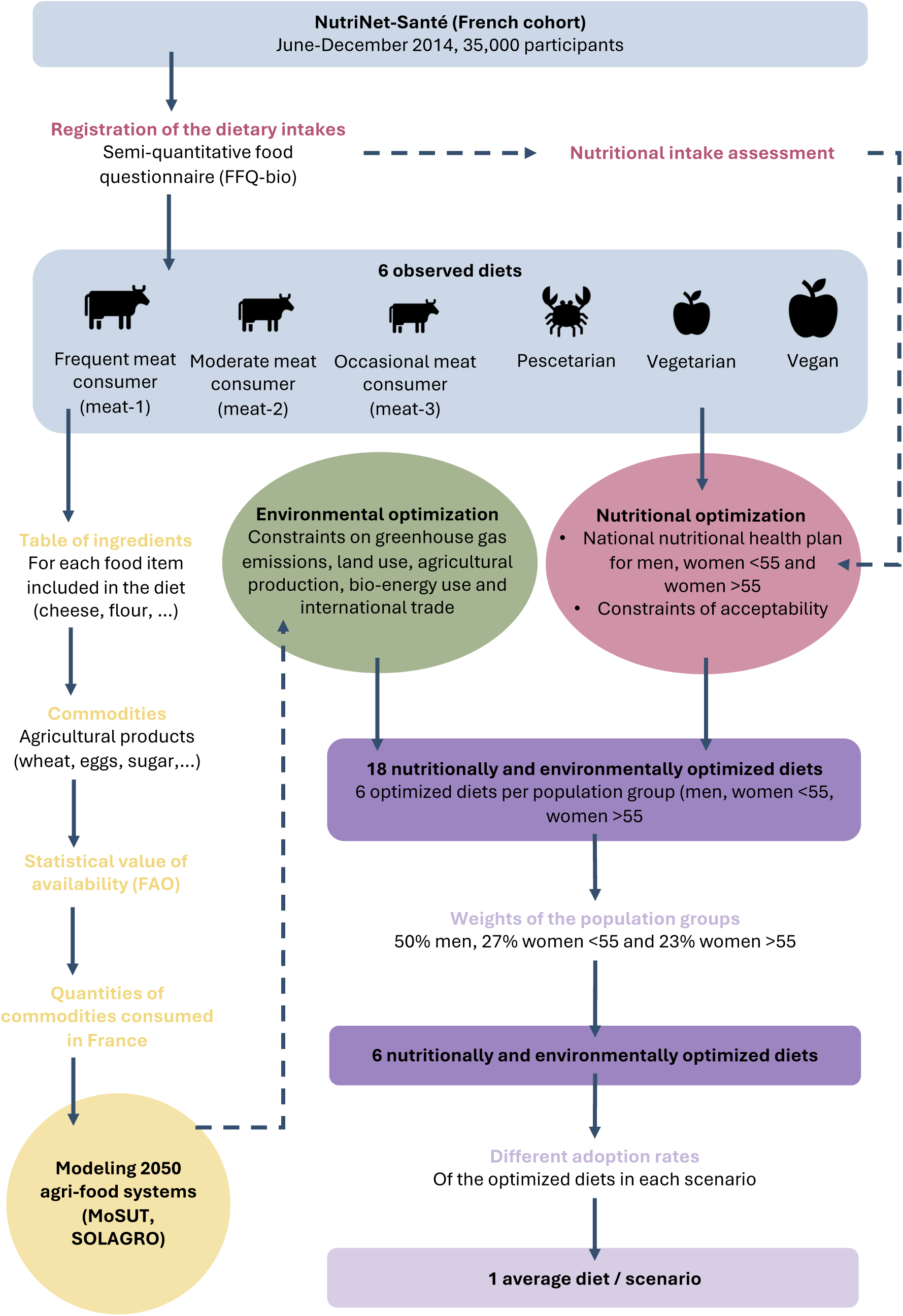
Construction of the 2050 scenarios’ diets

##### B Dietary shifts

Dietary shifts between 2025 (current observed diet) and 2050 (net-zero transition scenarios) were calculated with a cosine interpolation function, as follows:

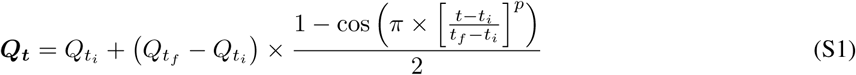

Where *Q_t_*is the quantity (given in grams per day per person) of a food group consumed in a year *t* (*t_i_* and *t_f_* refer to the initial and final years of dietary changes, set to 2025 and 2050 respectively). *p* is a parameter that modulates the shape of the curve, reflecting a more or less rapid change in diet. In the absence of evidence for either hypothesis, *p* was set to 1. Food consumption in the population was considered stationary for a 20-year period before 2025 and after 2050.

**Figure S2:**
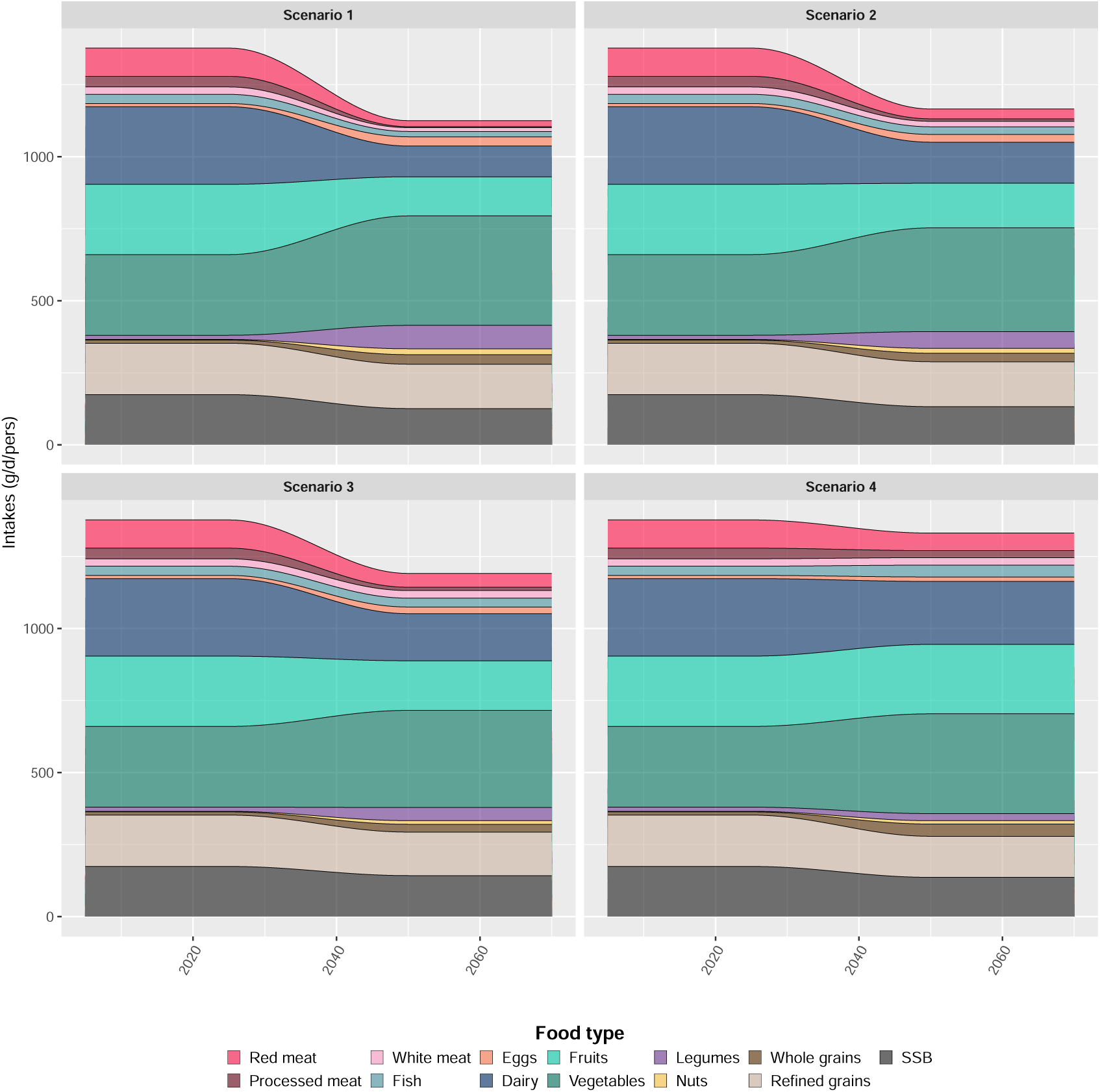
Projection over time of the French population average diet in the different net-zero transition scenarios.

**Figure S3:**
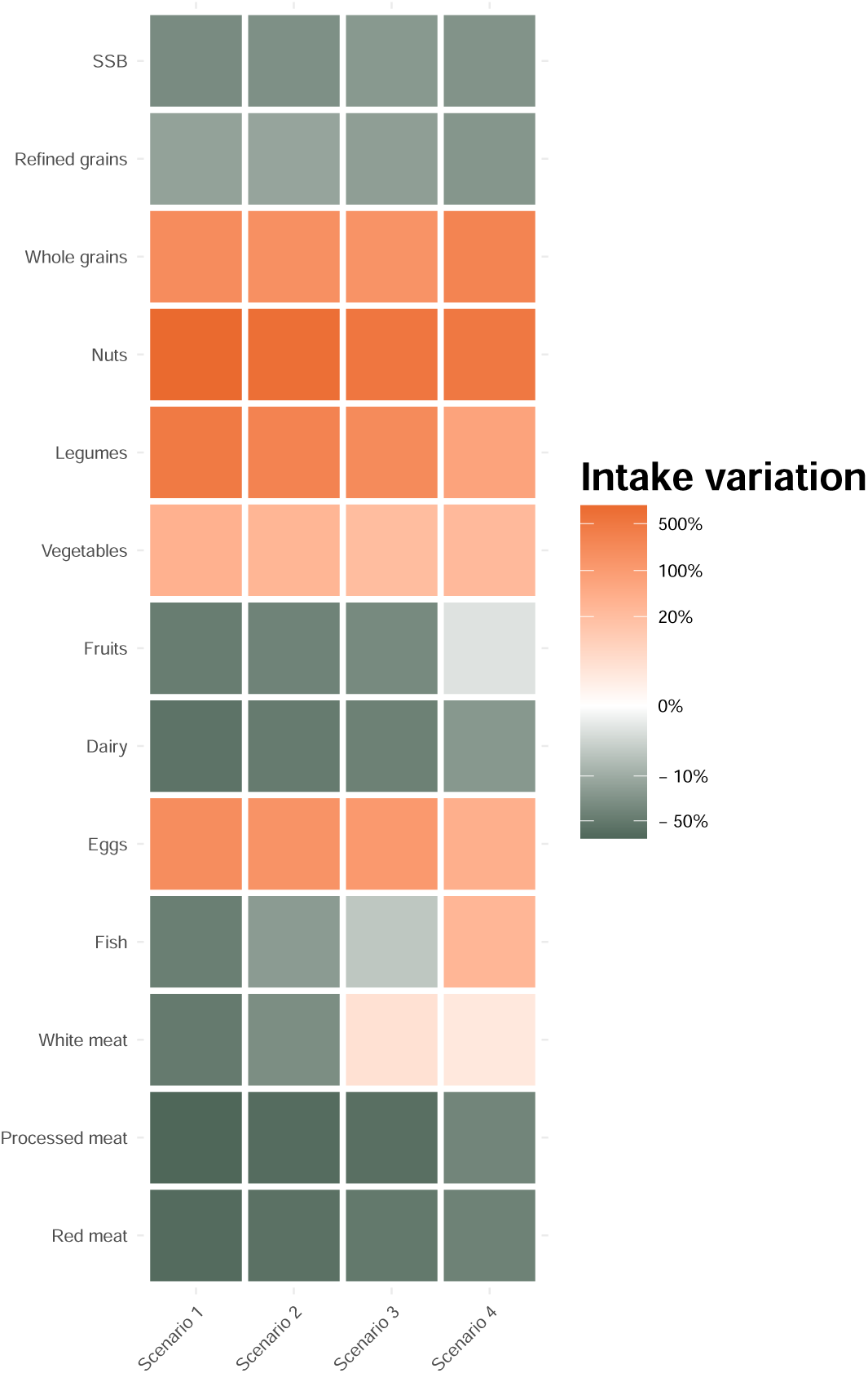
Variations in intakes in 2050 in different scenarios compared to the current observed average diet in France. SSB: Sugar-sweetened beverages.

##### C Selection of all-cause mortality risk meta-analyses

Meta-analyses on all-cause mortality risks associated with the consumption of food groups were taken from recent nutrition HIA publications,^18,19^ or have been updated from a recent umbrella review on the latest and most robust data.^26^ The certainty pertaining to the level of evidence was evaluated as “high” for whole grains and sugar-sweetened beverages, “moderate” for red meat, processed meat, fish, dairy, nuts and legumes, “low” for fruits, vegetables and refined grains, and “very low” for eggs and white meat.^18,20–25^ A sample of 1,000 values was generated for each extracted relative risk (RR) and its 95% confidence interval values, assuming a log-normal distribution. Using cubic spline interpolation, the dose-response relationships were reconstructed.

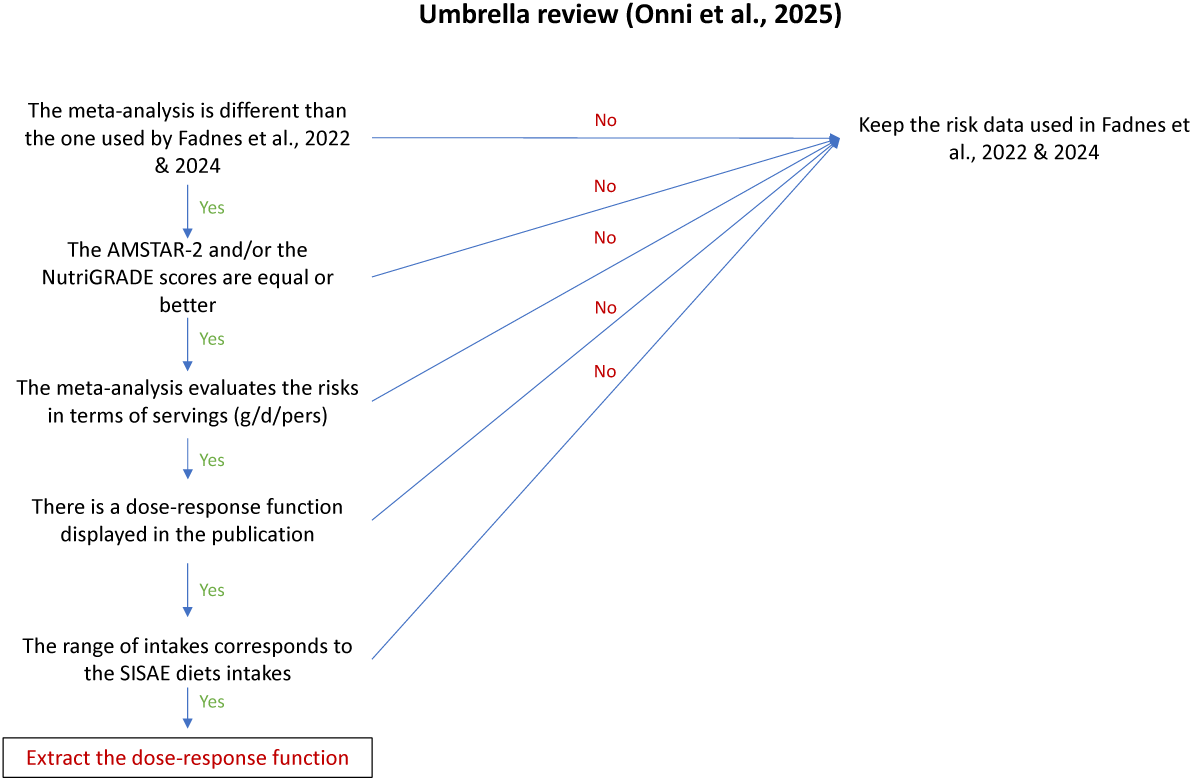

**Table S4:**
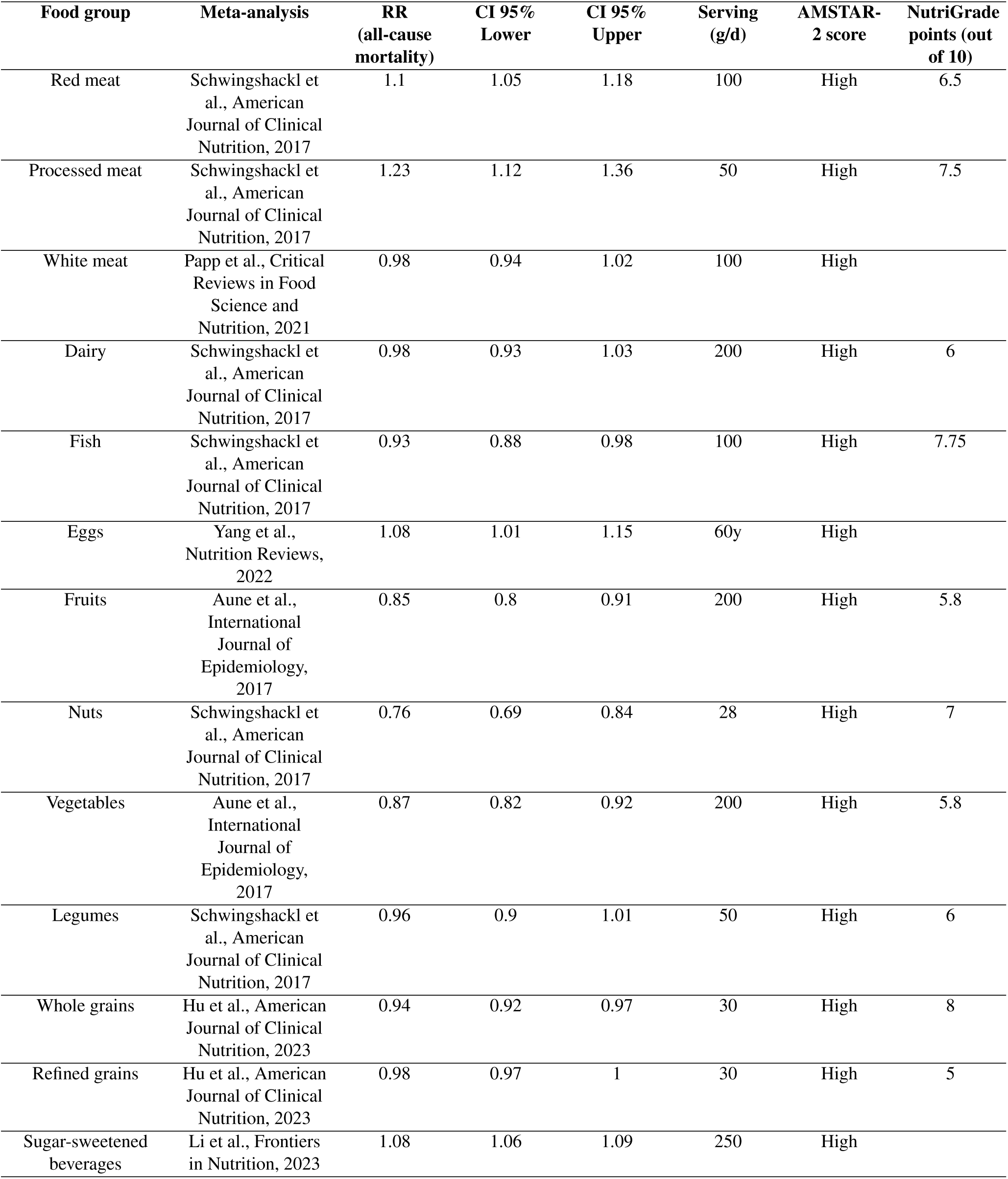
Relative risks (RR) of all-cause mortality related to the consumption of different food groups (with 95% confidence intervals, CI 95%, with the meta-analysis they were taken from, AMSTAR-2 and NutriGRADE scores

| Food group | Meta-analysis | RR<br>(all-cause<br>mortality) | CI 95%<br>Lower | CI 95%<br>Upper | Serving<br>(g/d) | AMSTAR-2 score | NutriGrade<br>points (out<br>of 10) |
| --- | --- | --- | --- | --- | --- | --- | --- |
| Red meat | Schwingshackl et al., American Journal of Clinical Nutrition, 2017 | 1.1 | 1.05 | 1.18 | 100 | High | 6.5 |
| Processed meat | Schwingshackl et al., American Journal of Clinical Nutrition, 2017 | 1.23 | 1.12 | 1.36 | 50 | High | 7.5 |
| White meat | Papp et al., Critical Reviews in Food Science and Nutrition, 2021 | 0.98 | 0.94 | 1.02 | 100 | High |  |
| Dairy | Schwingshackl et al., American Journal of Clinical Nutrition, 2017 | 0.98 | 0.93 | 1.03 | 200 | High | 6 |
| Fish | Schwingshackl et al., American Journal of Clinical Nutrition, 2017 | 0.93 | 0.88 | 0.98 | 100 | High | 7.75 |
| Eggs | Yang et al., Nutrition Reviews, 2022 | 1.08 | 1.01 | 1.15 | 60y | High |  |
| Fruits | Aune et al., International Journal of Epidemiology, 2017 | 0.85 | 0.8 | 0.91 | 200 | High | 5.8 |
| Nuts | Schwingshackl et al., American Journal of Clinical Nutrition, 2017 | 0.76 | 0.69 | 0.84 | 28 | High | 7 |
| Vegetables | Aune et al., International Journal of Epidemiology, 2017 | 0.87 | 0.82 | 0.92 | 200 | High | 5.8 |
| Legumes | Schwingshackl et al., American Journal of Clinical Nutrition, 2017 | 0.96 | 0.9 | 1.01 | 50 | High | 6 |
| Whole grains | Hu et al., American Journal of Clinical Nutrition, 2023 | 0.94 | 0.92 | 0.97 | 30 | High | 8 |
| Refined grains | Hu et al., American Journal of Clinical Nutrition, 2023 | 0.98 | 0.97 | 1 | 30 | High | 5 |
| Sugar-sweetened beverages | Li et al., Frontiers in Nutrition, 2023 | 1.08 | 1.06 | 1.09 | 250 | High |  |

##### D Attenuation in risk effect

In line with previous publications, the all-cause mortality risk of each food group included in the scenarios’ diets was reduced by a factor *m*, as follows:^18^

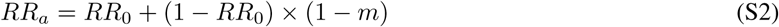

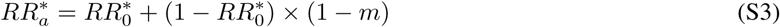

Where:

- *RR*_0_ is the RR extracted from the meta-analysis
- *RR_a_* is the adjusted RR
- *m* is a parameter of modulation of the RR value And *RR*^∗^_0_ and *RR*^∗^*_a_* are calculated as such:

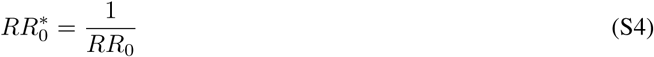

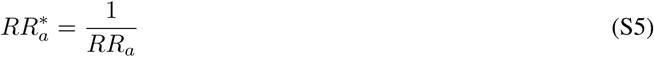

##### E Mortality risks calculation

The relative risk (RR) of the consumption of one food group *i* in a given year *t*, considering the contributions of the intakes of that food group in the past *x* years, is expressed as follows:

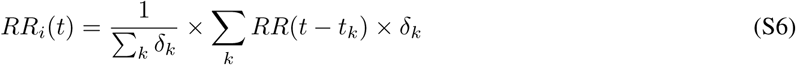

Where *i* is the food group, *t* is the year, *δ* is the weight attributed to each year after the food consumption, and *k* ∈ [[0*, x*]]

The RR associated with the complete diet in a given year *t* is simply expressed as the product of the RR of each food group *i* that makes up the diet:

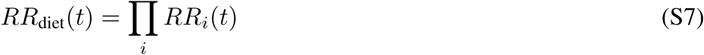

Based on the yearly projected value of life expectancy, we also calculated the gain in conditional life expectancy (LE), i.e., life expectancy if individuals have lived to a given age, and the years of life lost (YLL) averted by shifting diet. YLL prevented were calculated by summing the number of age-specific prevented deaths yearly in each scenario, multiplied by the number of remaining years to live, calculated as the difference between conditional LE and the modeled age at death.

We took care of reporting the prevented deaths in year *t* and age category *a* to year *t* + 1 and age category *a* + 1.

##### F Food groups contributions

Contributions of each food group to the health impact results have been extracted from the calculation of the number of deaths prevented.

In a given scenario, the age-and-year-specific number of deaths prevented was calculated as follows:

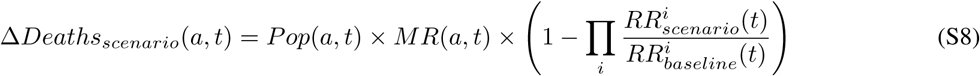

If we’re interested in a specific food group *j*, here’s how it appears in the same equation:

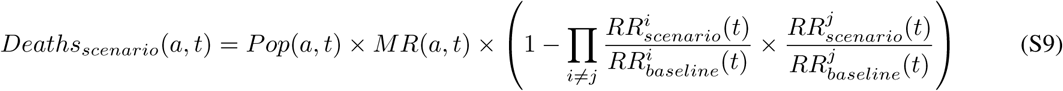

The fraction of the two relative risks (RR) will take the following form, ∀*δ^j^_scenario_* ∈ ℜ

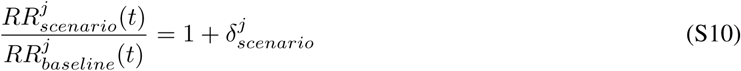

Thus, the contribution is calculated as follows:

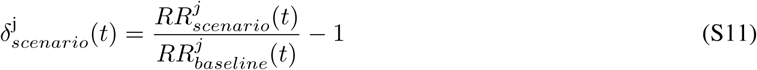

##### G Sensitivity analyses

The main analysis assumed a cosine interpolation implementation of diets from 2025 to 2050, as well as a 10-year linear time to full effect and a 25% reduction in mortality risk for each food group. Different sensitivity analyses were performed. The first one explored a transition in diet starting in 2016, right after the Paris Agreement; the second one explored an immediate dietary implementation in 2025; third, we conducted analyses with no attenuation or a 50% attenuation in the RRs. Afterwards, we used either an immediate or a 20-year-long time to full effect. Lastly, we considered a stable population over time, where the 2025 French age structure was maintained.

The number of prevented deaths and the percentage of deviation from the main analysis for different analyses are displayed in the following tables.

**Table S5:**
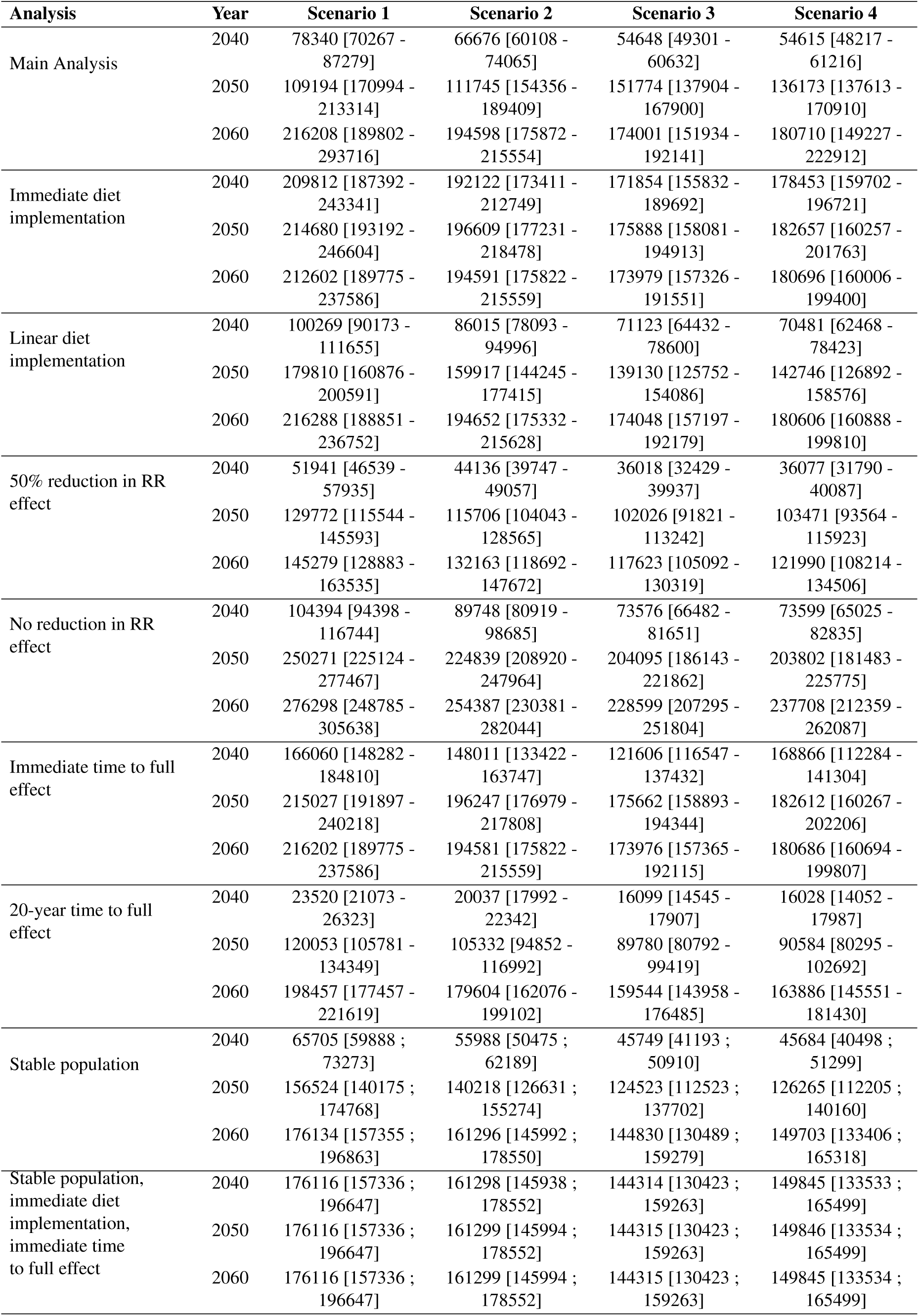
Number of deaths prevented in main and sensitivity analyses.

| Analysis | Year | Scenario 1 | Scenario 2 | Scenario 3 | Scenario 4 |
| --- | --- | --- | --- | --- | --- |
| Main Analysis | 2040 | 78340 [70267 - 87279] | 66676 [60108 - 74065] | 54648 [49301 - 60632] | 54615 [48217 - 61216] |
|  | 2050 | 109194 [170994 - 213314] | 111745 [154356 - 189409] | 151774 [137904 - 167900] | 136173 [137613 - 170910] |
|  | 2060 | 216208 [189802 - 293716] | 194598 [175872 - 215554] | 174001 [151934 - 192141] | 180710 [149227 - 222912] |
| Immediate diet implementation | 2040 | 209812 [187392 - 243341] | 192122 [173411 - 212749] | 171854 [155832 - 189692] | 178453 [159702 - 196721] |
|  | 2050 | 214680 [193192 - 246604] | 196609 [177231 - 218478] | 175888 [158081 - 194913] | 182657 [160257 - 201763] |
|  | 2060 | 212602 [189775 - 237586] | 194591 [175822 - 215559] | 173979 [157326 - 191551] | 180696 [160006 - 199400] |
| Linear diet implementation | 2040 | 100269 [90173 - 111655] | 86015 [78093 - 94996] | 71123 [64432 - 78600] | 70481 [62468 - 78423] |
|  | 2050 | 179810 [160876 - 200591] | 159917 [144245 - 177415] | 139130 [125752 - 154086] | 142746 [126892 - 158576] |
|  | 2060 | 216288 [188851 - 236752] | 194652 [175332 - 215628] | 174048 [157197 - 192179] | 180606 [160888 - 199810] |
| 50% reduction in RR effect | 2040 | 51941 [46539 - 57935] | 44136 [39747 - 49057] | 36018 [32429 - 39937] | 36077 [31790 - 40087] |
|  | 2050 | 129772 [115544 - 145593] | 115706 [104043 - 128565] | 102026 [91821 - 113242] | 103471 [93564 - 115923] |
|  | 2060 | 145279 [128883 - 163535] | 132163 [118692 - 147672] | 117623 [105092 - 130319] | 121990 [108214 - 134506] |
| No reduction in RR effect | 2040 | 104394 [94398 - 116744] | 89748 [80919 - 98685] | 73576 [66482 - 81651] | 73599 [65025 - 82835] |
|  | 2050 | 250271 [225124 - 277467] | 224839 [208920 - 247964] | 204095 [186143 - 221862] | 203802 [181483 - 225775] |
|  | 2060 | 276298 [248785 - 305638] | 254387 [230381 - 282044] | 228599 [207295 - 251804] | 237708 [212359 - 262087] |
| Immediate time to full effect | 2040 | 166060 [148282 - 184810] | 148011 [133422 - 163747] | 121606 [116547 - 137432] | 168866 [112284 - 141304] |
|  | 2050 | 215027 [191897 - 240218] | 196247 [176979 - 217808] | 175662 [158893 - 194344] | 182612 [160267 - 202206] |
|  | 2060 | 216202 [189775 - 237586] | 194581 [175822 - 215559] | 173976 [157365 - 192115] | 180686 [160694 - 199807] |
| 20-year time to full effect | 2040 | 23520 [21073 - 26323] | 20037 [17992 - 22342] | 16099 [14545 - 17907] | 16028 [14052 - 17987] |
|  | 2050 | 120053 [105781 - 134349] | 105332 [94852 - 116992] | 89780 [80792 - 99419] | 90584 [80295 - 102692] |
|  | 2060 | 198457 [177457 - 221619] | 179604 [162076 - 199102] | 159544 [143958 - 176485] | 163886 [145551 - 181430] |
| Stable population | 2040 | 65705 [59888 ; 73273] | 55988 [50475 ; 62189] | 45749 [41193 ; 50910] | 45684 [40498 ; 51299] |
|  | 2050 | 156524 [140175 ; 174768] | 140218 [126631 ; 155274] | 124523 [112523 ; 137702] | 126265 [112205 ; 140160] |
|  | 2060 | 176134 [157355 ; 196863] | 161296 [145992 ; 178550] | 144830 [130489 ; 159279] | 149703 [133406 ; 165318] |
| Stable population, immediate diet implementation, immediate time to full effect | 2040 | 176116 [157336 ; 196647] | 161298 [145938 ; 178552] | 144314 [130423 ; 159263] | 149845 [133533 ; 165499] |
|  | 2050 | 176116 [157336 ; 196647] | 161299 [145994 ; 178552] | 144315 [130423 ; 159263] | 149846 [133534 ; 165499] |
|  | 2060 | 176116 [157336 ; 196647] | 161299 [145994 ; 178552] | 144315 [130423 ; 159263] | 149845 [133534 ; 165499] |

**Table S6:** Percentage of change in the number of deaths prevented in different analyses compared to the main one.

| Analysis | Year | Scenario 1 | Scenario 2 | Scenario 3 | Scenario 4 |
| --- | --- | --- | --- | --- | --- |
| Immediate diet implementation | 2040 | 112.0% [111; 113.4] | 122.0% [123.5; 121.1] | 136.3% [136.4; 136.5] | 132.3% [132.9; 131] |
|  | 2050 | 12.6% [12.3; 12.6] | 14.7% [14.7; 15] | 15.7% [15.9; 15.7] | 18.5% [18.9; 18.2] |
|  | 2060 | 0.0% [0; 0] | 0.0% [0; 0] | 0.0% [0; 0] | 0.0% [0; 0] |
| 20-year time to full effect | 2040 | -70.0% [-70.1; -69.8] | -69.9% [-70.1; -69.8] | -70.7% [-70.7; -70.5] | -70.7% [-70.9; -70.6] |
|  | 2050 | -37.0% [-37.7; -37] | -38.5% [-38.6; -38.3] | -40.8% [-40.9; -40.8] | -41.2% [-41.3; -41] |
|  | 2060 | -6.7% [-6.5; -6.7] | -7.7% [-7.5; -7.6] | -8.3% [-8.4; -8.1] | -9.3% [-9.6; -9.2] |
| No reduction in RR effect | 2040 | 33.9% [34.3%; 33.8] | 34.6% [34.6%; 34.6] | 34.8% [34.8%; 34.7] | 34.8% [34.9%; 34.6] |
|  | 2050 | 31.1% [31.7%; 30.1] | 31.4% [32%; 30.9] | 32.1% [32.5%; 31.8] | 32.3% [32.7%; 32.1] |
|  | 2060 | 29.0% [30%; 29] | 30.7% [31.4%; 30] | 31.4% [31.9%; 31.1] | 31.5% [32%; 31.2] |
| 50% reduction in RR effect | 2040 | -33.7% [-33.8%; -33.6] | -33.8% [-33.9%; -33.8] | -34.1% [-34.2%; -34] | -33.9% [-34.1%; -34] |
|  | 2050 | -32.0% [-32.4%; -31.7] | -32.4% [-32.6%; -32.1] | -32.8% [-33%; -32.6] | -32.8% [-33%; -32.7] |
|  | 2060 | -31.7% [-32.1%; -31.2] | -32.1% [-32.3%; -31.8] | -32.4% [-32.6%; -32.2] | -32.5% [-32.9%; -32.3] |
| Immediate diet implementation | 2040 | 167.8% [166.7; 168.5] | 188.1% [189.7; 187.2] | 214.5% [215%; 212.9] | 226.7% [229.7%; 222] |
|  | 2050 | 12.5% [12.2; 12.5] | 14.9% [14.8; 15] | 15.9% [15.9; 15.7] | 18.0% [19; 18.1] |
|  | 2060 | 0.0% [0; 0] | 0.0% [0; 0] | 0.0% [0; 0] | 0.0% [0; 0] |
| Linear diet implementation | 2040 | 28.0% [28.3; 27.9] | 29.0% [29.9; 28.3] | 30.1% [30.7; 29.6] | 29.0% [29.5; 28.1] |
|  | 2050 | -5.8% [-5.9; -6.6] | -6.6% [-6.6; -6.3] | -8.3% [-8.3; -8.2] | -7.3% [-7.1%; -7.2] |
|  | 2060 | 0.0% [0; 0] | 0.0% [0; 0] | 0.0% [0; 0] | -0.1% [0.1%; 0] |
| Stable population | 2040 | -16.1% [-16.1 ; -16] | -16.0% [-16 ; 16] | -16.3% [16.4 ; -16] | -16.0% [-16 ; 16.2] |
|  | 2050 | -18.0% [-18 ; -18.1] | -18.1% [-18 ; 18] | -18.0% [-17.9 ; -18] | -18.0% [-17.9 ; -18] |
|  | 2060 | -17.2% [-17.1 ; -17.2] | -17.1% [-17.1 ; -17.2] | -17.1% [-17.7 ; -17.1] | -17.2% [-17.1 ; -17.1] |
| Stable population, immediate diet implementation, immediate time to full effect | 2040 | 124.8% [123.9 ; 125.3] | 141.9% [141.9 ; 141.1] | 164.1% [164.5 ; 162.7] | 174.4% [170.9 ; 170.4] |
|  | 2050 | -7.8% [-7.9 ; -7.8] | -5.8% [-5.8 ; -5.7] | -4.9% [-4.9 ; -5.1] | -2.7% [-2.3 ; 3.2] |
|  | 2060 | -17.2% [-17.1 ; -17.2] | -17.1% [-17 ; -17.2] | -17.1% [-17.7 ; -17.1] | -17.1% [-17.7 ; -17.1] |

**Figure S4:**
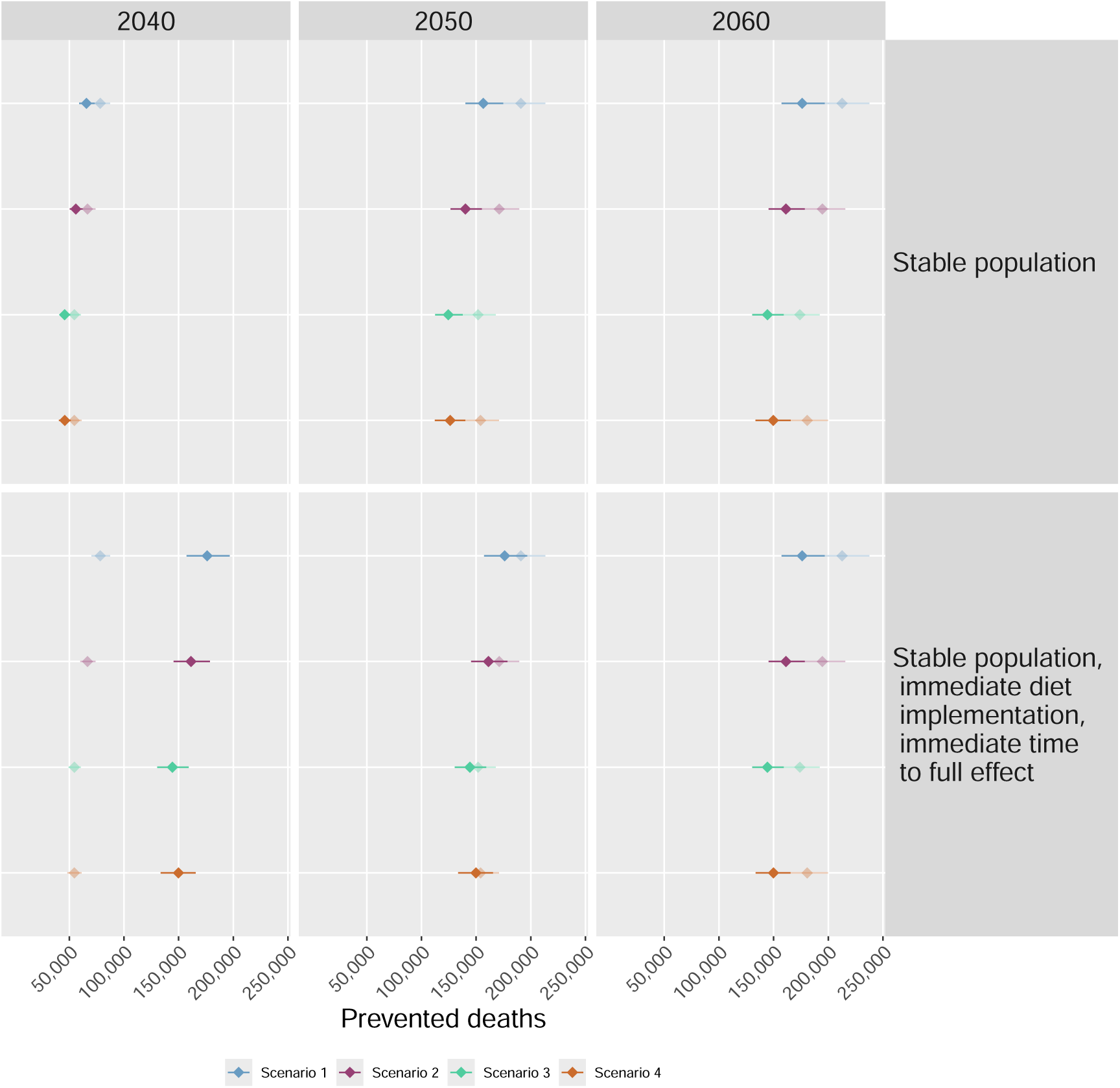
Number of deaths prevented in 2040, 2050 and 2060 under different assumptions. The points in transparency represent the main analysis results.

##### H Current burden of poor dietary habits in France

In order to validate our health impact assessment model and estimate the current burden of poor diets in France, we conducted an additional analysis in which we applied an optimized diet to the 2025 French adult population and computed the health benefits in terms of all-cause mortality compared to the consumption of the currently observed average diet.^14^ There is no scientific consensus on the diet that would bring the most health benefits. We chose to apply the optimized diet designed by Fadnes et al., minimizing the all-cause mortality risks for the intakes of each of the thirteen food groups included (which are the same as the ones included in the diets studied in this paper).^19^ We also conducted this analysis with the four 2050 diets that were analyzed in this study. As we aimed to assess the current burden of poor diets, we considered an immediate implementation of diets with no time lag in the health impacts. All the other assumptions were the same as in the main analysis.

**Table S7:** Average gain in years of life expectancy (LE) and number of deaths prevented in 2025 in the French population through different dietary shifts as compared to maintaining the currently observed diet. Implementation of diets and the time lag of mortality benefits were considered to be immediate

| Scenario | LE | Prevented deaths |
| --- | --- | --- |
| Optimized diet | 6.3 [5.2 - 7.3] | 392,800 [349,725 - 431,537] |
| Scenario 1 | 2.3 [2.1 - 2.7] | 189,098 [169,408 - 210,534] |
| Scenario 2 | 2.1 [1.9 - 2.4] | 173,563 [156,818 - 191,664] |
| Scenario 3 | 1.9 [1.7 - 2.1] | 155,669 [140,974 - 171,435] |
| Scenario 4 | 1.9 [1.7 - 2.2] | 161,505 [144,271 - 177,987] |

**Figure S5:**
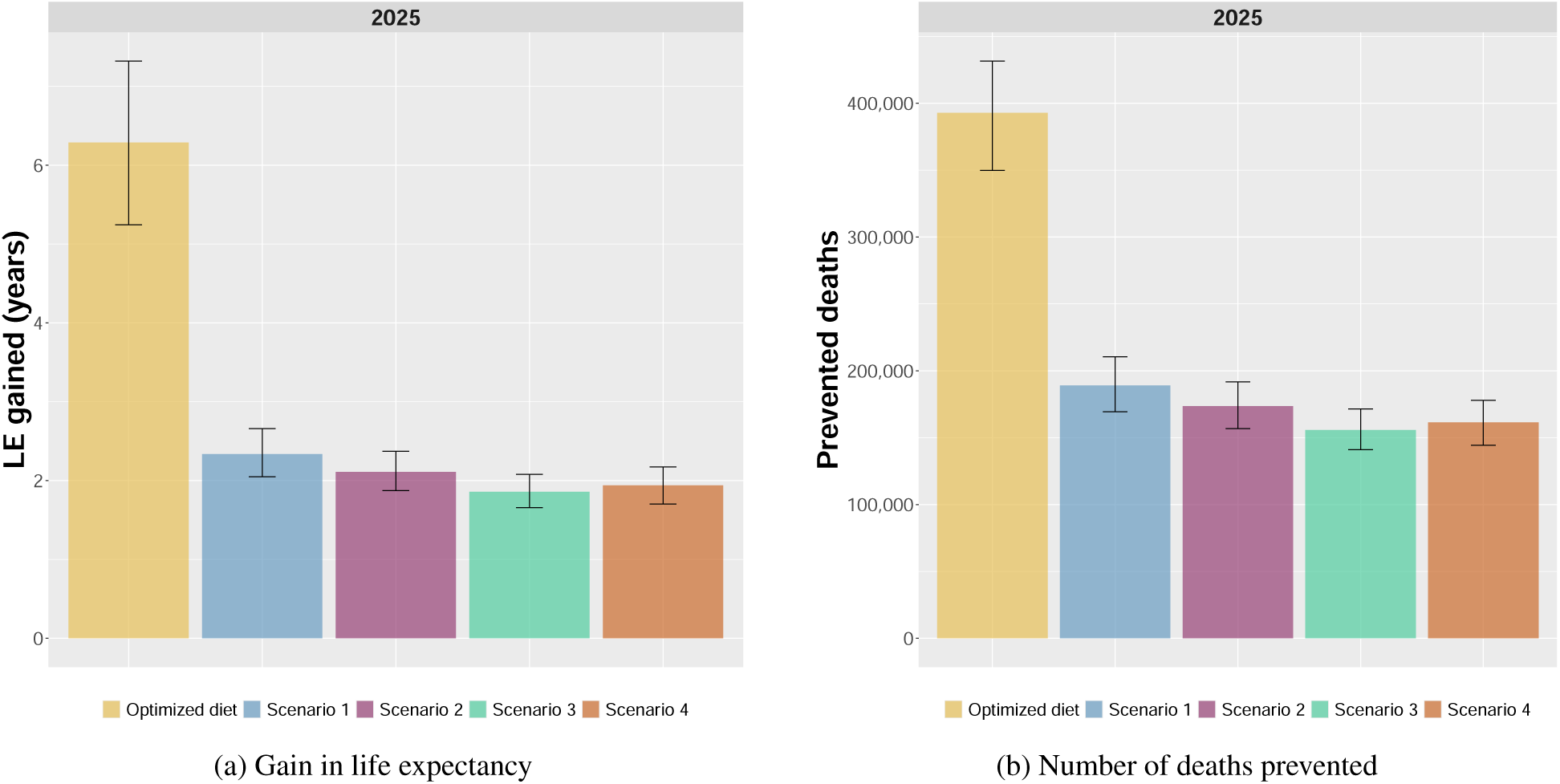
Average gains in years of life expectancy (a) and number of deaths prevented (b) in 2025 through various dietary shifts

**Table S8:** Gains in years of life expectancy in 2025 through various dietary shifts for people aged 20, 40, 60 and 80.

| Scenario | LE |  |  |  |
| --- | --- | --- | --- | --- |
|  | 20 | 40 | 60 | 80 |
| Optimized diet | 8.5 [7.2 - 9.9] | 8.2 [6.9 - 9.5] | 7.4 [6.1 - 8.6] | 5.3 [4.4 - 6.2] |
| Scenario 1 | 3.3 [2.9 - 3.7] | 3.1 [2.7 - 3.6] | 2.7 [2.4 - 3.1] | 1.9 [1.6 - 2.1] |
| Scenario 2 | 3.0 [2.6 - 3.3] | 2.8 [2.5 - 3.2] | 2.5 [2.2 - 2.8] | 1.7 [1.5 - 1.9] |
| Scenario 3 | 2.6 [2.3 - 2.9] | 2.5 [2.2 - 2.8] | 2.2 [1.9 - 2.4] | 1.5 [1.3 - 1.6] |
| Scenario 4 | 2.7 [2.4 - 3.0] | 2.6 [2.3 - 2.9] | 2.3 [2.0 - 2.6] | 1.5 [1.3 - 1.7] |

**Figure S6:**
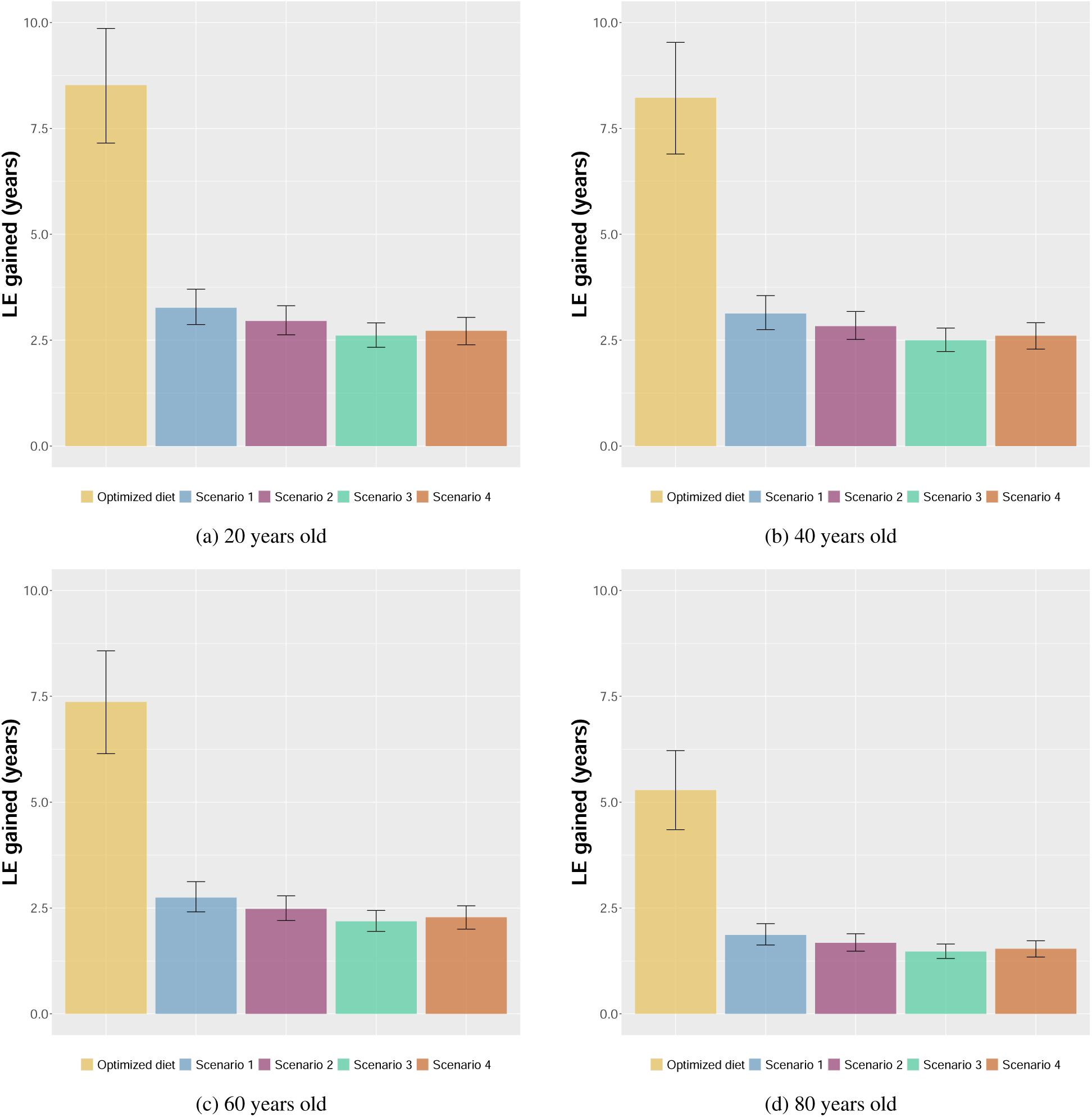
Gains in years of life expectancy in 2025 through various dietary shifts for people aged 20 (a), 40 (b), 60 and 80 (d)

In 2025, if the French adult population had eaten one of the four 2050 diets instead of the currently observed one, it could have prevented up to 189,000 [169,000 - 211,000] deaths with the diet of scenario 1, corresponding to an average gain in life expectancy of 2.3 [2.1 - 2.7] years. In comparison, if the population had consumed the optimized diet designed by Fadnes et al., it could have prevented 393,000 [350,000 - 432,000] deaths, corresponding to an average increase in life expectancy of 6.3 [5.2 - 7.3] years (Table S7, Figure S5). This indicates that the current burden of poor diets could be very important in the French population, with about 61% [55 - 67%] of mortality that could have been prevented in 2025. Our results are consistent with previous studies. In particular, Laine et al., showed that a high adherence to the 2019 Planetary Health Diet^52^ could prevent up to 63% of deaths in a European population.^53^ To go further into details, we computed the gain in life expectancy at the age of 20, 40, 60, and 80 with these dietary shifts. The consumption of the optimized diet could have increased life expectancy by between 5.3 [4.4 - 6.2] years at 80 years old and 8.5 [7.2 - 9.9] at 20 years old (Table S8, Figure S6). Our results are in the same order of magnitude as the study of Fadnes et al. (up to 8.1 [6.7 - 9.4] years and 9.3 [7.8 - 10.8] years of gain in life expectancy for 20-year-old French females and males respectively).^19^ Overall, these results show that there is potential for more health gains thanks to dietary shifts, compared to the net-zero emission scenarios that were analyzed in this study. Nevertheless, the optimized diet designed by Fadnes et al. implies drastic changes in dietary habits and could be considered as an upper limit in the health benefits that we could expect from nutritional aspects.

## References

[1] Rockström J, Thilsted SH, Willett WC, Gordon LJ, Herrero M, Hicks CC, et al. The EAT–Lancet Commission on healthy, sustainable, and just food systems. The Lancet. 2025 10;406:1625–700. Available from: https://linkinghub.elsevier.com/retrieve/pii/S0140673625012012.

[2] Crippa M, Solazzo E, Guizzardi D, Monforti-Ferrario F, Tubiello FN, Leip A. Food systems are responsible for a third of global anthropogenic GHG emissions. Nature Food. 2021 3;2:198–209. Available from: https://www.nature.com/articles/s43016-021-00225-9.

[3] Midler E, Voirin S, Hammani A, Sgambati E, Kasriel Y, Mondon S, et al. Accélérer la transition climatique avec un système alimentaire bas carbone, résilient et juste. Haut Conseil pour le Climat; 2024. Available from: https://www.hautconseilclimat.fr/publications/accelerer-la-transition-climatique-avec-un-systeme-alimentaire-bas-carbone-resilient-et-juste/.

[4] Poore J, Nemecek T. Reducing food’s environmental impacts through producers and consumers. Science. 2018 6;360:987–92. Available from: https://pubmed.ncbi.nlm.nih.gov/29853680/.

[5] HLPE. Nutrition and food systems. A report by the High Level Panel of Experts on Food Security and Nutrition of the Committee on World Food Security, Rome. 2017. Available from: www.fao.org/cfs/cfs-hlpe.

[6] Hay SI, Ong KL, Santomauro DF, A B, Aalipour MA, Aalruz H, et al. Burden of 375 diseases and injuries, risk-attributable burden of 88 risk factors, and healthy life expectancy in 204 countries and territories, including 660 subnational locations, 1990–2023: a systematic analysis for the Global Burden of Disease Study 2023. The Lancet. 2025 10;406:1873–922. Available from: https://www.thelancet.com/action/showFullText?pii=S014067362501637Xhttps://www.thelancet.com/action/showAbstract?pii=S014067362501637Xhttps://www.thelancet.com/journals/lancet/article/PIIS0140-6736(25)01637-X/abstract.

[7] Kesse-Guyot E, Chayre A, Perraud E, Berger S, Richard A, Berlivet J, et al. Association between dietary environmental pressures and major chronic diseases: assessment from the prospective NutriNet-Santé cohort. The Lancet Regional Health -Europe. 2025 12;59:101481. Available from: https://pubmed.ncbi.nlm.nih.gov/41127057/.

[8] Hamilton I, Kennard H, McGushin A, Höglund-Isaksson L, Kiesewetter G, Lott M, et al. The public health implications of the Paris Agreement: a modelling study. The Lancet Planetary Health. 2021 2;5:e74–83. Available from: https://linkinghub.elsevier.com/retrieve/pii/S2542519620302497.

[9] Milner J, Turner G, Ibbetson A, Colombo PE, Green R, Dangour AD, et al. Impact on mortality of pathways to net zero greenhouse gas emissions in England and Wales: a multisectoral modelling study. The Lancet Planetary Health. 2023;7:e128–36. Available from: 10.1016/S2542-5196(22)00310-2.

[10] Moutet L, Bernard P, Green R, Milner J, Haines A, Slama R, et al. The public health co-benefits of strategies consistent with net-zero emissions: a systematic review. The Lancet Planetary Health. 2025 2;9:e145–56. Available from: https://linkinghub.elsevier.com/retrieve/pii/S2542519624003309.

[11] Barbier C, Couturier C, Dumas P, Kesse-Guyot E, Baudry J, Pharabod I, et al. Simulation prospective du Système Alimentaire et de son Empreinte énergétique et carbone. 2022. Available from: https://solagro.org.

[12] ADEME. Prospective - Transition(s) 2050 - Summary; 2021. Available from: https://librairie.ademe.fr.

[13] Hess JJ, Ranadive N, Boyer C, Aleksandrowicz L, Anenberg SC, Aunan K, et al. Guidelines for Modeling and Reporting Health Effects of Climate Change Mitigation Actions. Environmental Health Perspectives. 2020 11;128:1–10. Available from: https://ehp.niehs.nih.gov/doi/10.1289/EHP6745.

[14] Baudry J, Allés B, Péneau S, Touvier M, Méjean C, Hercberg S, et al. Dietary intakes and diet quality according to levels of organic food consumption by French adults: cross-sectional findings from the NutriNet-Santé Cohort Study. Public Health Nutrition. 2016 3;20:638–48. Available from: https://www.cambridge.org/core/product/identifier/S1368980016002718/type/journal_article.

[15] Kesse-Guyot E, Lairon D, Allès B, Seconda L, Rebouillat P, Brunin J, et al. Key Findings of the French BioNutriNet Project on Organic Food–Based Diets: Description, Determinants, and Relationships to Health and the Environment. Advances in Nutrition. 2022 1;13:208–24. Available from: https://linkinghub.elsevier.com/retrieve/pii/S2161831322005373.

[16] Kesse-Guyot E, Allès B, Brunin J, Fouillet H, Dussiot A, Mariotti F, et al. Nutritionally adequate and environmentally respectful diets are possible for different diet groups: an optimized study from the NutriNet-Santé cohort. The American Journal of Clinical Nutrition. 2022 12;116:1621–33. Available from: https://linkinghub.elsevier.com/retrieve/pii/S000291652303695X.

[17] ANSES. Actualisation des repères du PNNS : révision des repères de consommations alimentaires; 2016. Available from: https://www.anses.fr.

[18] Fadnes LT, Økland JM, Øystein A Haaland, Johansson KA. Estimating impact of food choices on life expectancy: A modeling study. PLoS Medicine. 2022 2;19:e1003889. Available from: https://journals.plos.org/plosmedicine/article?id=10.1371/journal.pmed.1003889.

[19] Fadnes LT, Arjmand EJ, Økland JM, Celis-Morales C, Livingstone KM, Balakrishna R, et al. Life expectancy gains from dietary modifications: a comparative modeling study in 7 countries. The American Journal of Clinical Nutrition. 2024 7;120:170–7. Available from: https://linkinghub.elsevier.com/retrieve/pii/S0002916524004544.

[20] Papp RE, Hasenegger V, Ekmekcioglu C, Schwingshackl L. Association of poultry consumption with cardiovascular diseases and all-cause mortality: a systematic review and dose response meta-analysis of prospective cohort studies. Critical Reviews in Food Science and Nutrition. 2023 6;63:2366–87. Available from: https://www.tandfonline.com/doi/full/ 10.1080/10408398.2021.1975092.

[21] Yang PF, Wang CR, Hao FB, Peng Y, Wu JJ, Sun WP, et al.. Egg consumption and risks of all-cause and cause-specific mortality: A dose-response meta-analysis of prospective cohort studies; 2022. Available from: 10.1093/nutrit/nuac002.

[22] Hu H, Zhao Y, Feng Y, Yang X, Li Y, Wu Y, et al. Consumption of whole grains and refined grains and associated risk of cardiovascular disease events and all-cause mortality: a systematic review and dose-response meta-analysis of prospective cohort studies. American Journal of Clinical Nutrition. 2023;117:149–59. Available from: 10.1016/j.ajcnut.2022.10.010.

[23] Li B, Yan N, Jiang H, Cui M, Wu M, Wang L, et al. Consumption of sugar sweetened beverages, artificially sweetened beverages and fruit juices and risk of type 2 diabetes, hypertension, cardiovascular disease, and mortality: A meta-analysis. Frontiers in Nutrition. 2023 3;10:1019534. Available from: https://www.frontiersin.org/articles/10.3389/fnut.2023.1019534/full.

[24] Aune D, Giovannucci E, Boffetta P, Fadnes LT, Keum NN, Norat T, et al. Fruit and vegetable intake and the risk of cardiovascular disease, total cancer and all-cause mortality-A systematic review and dose-response meta-analysis of prospective studies. International Journal of Epidemiology. 2017;46:1029–56. Available from: https://academic.oup.com/ije/article/46/3/1029/3039477.

[25] Schwingshackl L, Schwedhelm C, Hoffmann G, Lampousi AM, Knüppel S, Iqbal K, et al. Food groups and risk of all-cause mortality: a systematic review and meta-analysis of prospective studies ,. The American Journal of Clinical Nutrition. 2017 6;105:1462–73. Available from: https://linkinghub.elsevier.com/retrieve/pii/S0002916522049206.

[26] Onni AT, Balakrishna R, Perillo M, Amato M, Arjmand EJ, Thomassen LM, et al. Umbrella review of systematic reviews and meta-analyses on consumption of different food groups and risk of all-cause mortality. Advances in Nutrition. 2025 2;0:100393. Available from: https://advances.nutrition.org/action/showFullText?pii=S2161831325000298https://advances.nutrition.org/action/showAbstract?pii=S2161831325000298https://advances.nutrition.org/article/S2161-8313(25)00029-8/abstract.

[27] Shea BJ, Reeves BC, Wells G, Thuku M, Hamel C, Moran J, et al. AMSTAR 2: A critical appraisal tool for systematic reviews that include randomised or non-randomised studies of healthcare interventions, or both. BMJ (Online). 2017 9;358. Available from: https://www.bmj.com/content/358/bmj.j4008https://www.bmj.com/content/358/bmj.j4008.abstract.

[28] Schwingshackl L, Knüppel S, Schwedhelm C, Hoffmann G, Missbach B, Stelmach-Mardas M, et al. Perspective: NutriGrade: A scoring system to assess and judge the meta-evidence of randomized controlled trials and cohort studies in nutrition research. Advances in Nutrition. 2016;7:994–1004. Available from: http://advances.nutrition.org.

[29] Afshin A, Sur PJ, Fay KA, Cornaby L, Ferrara G, Salama JS, et al. Health effects of dietary risks in 195 countries, 1990–2017: a systematic analysis for the Global Burden of Disease Study 2017. The Lancet. 2019 5;393:1958–72. Available from: https://linkinghub.elsevier.com/retrieve/pii/S0140673619300418.

[30] INSEE. Résultats détaillés des projections de population 2021-2070 pour la France – Scénario central; 2021. Available from: https://www.insee.fr/fr/statistiques/5894083?sommaire=5760764.

[31] Colombo PE, Milner J, Scheelbeek PF, Taylor A, Parlesak A, Kastner T, et al. Pathways to “5-a-day”: modeling the health impacts and environmental footprints of meeting the target for fruit and vegetable intake in the United Kingdom. The American Journal of Clinical Nutrition. 2021 8;114:530–9. Available from: https://linkinghub.elsevier.com/retrieve/pii/S0002916522003689.

[32] Milner J, Green R, Dangour AD, Haines A, Chalabi Z, Spadaro J, et al. Health effects of adopting low greenhouse gas emission diets in the UK. BMJ Open. 2015;5.

[33] Kesse-Guyot E, Chaltiel D, Wang J, Pointereau P, Langevin B, Allès B, et al. Sustainability analysis of French dietary guidelines using multiple criteria. Nature Sustainability. 2020 3;3:377–85. Available from: https://www.nature.com/articles/s41893-020-0495-8.

[34] Kesse-Guyot E, Baudry J, Berlivet J, Perraud E, Julia C, Touvier M, et al. To be climate-friendly, food-based dietary guidelines must include limits on total meat consumption – modeling from the case of France. International Journal of Behavioral Nutrition and Physical Activity 2025 22:1. 2025 7;22:1-15. Available from: https://ijbnpa.biomedcentral.com/articles/10.1186/s12966-025-01786-9.

[35] Hu FB. Dietary pattern analysis: a new direction in nutritional epidemiology. Current Opinion in Lipidology. 2002 2;13:3–9. Available from: http://journals.lww.com/00041433-200202000-00002.

[36] Eker S, Reese G, Obersteiner M. Modelling the drivers of a widespread shift to sustainable diets. Nature Sustainability. 2019 7;2:725–35. Available from: https://www.nature.com/articles/s41893-019-0331-1.

[37] Lee PN, Forey BA, Coombs KJ. Systematic review with meta-analysis of the epidemiological evidence in the 1900s relating smoking to lung cancer. BMC Cancer. 2012 9;12:1–90. Available from: https://bmccancer.biomedcentral.com/articles/10.1186/1471-2407-12-385.

[38] Anderson R, Armstrong B, Cullinan P, Derwent RG, Donaldson K, Harrison R, et al. The Mortality Effects of Long-Term Exposure to Particulate Air Pollution in the United Kingdom: A report by the Committee on the Medical Effects of Air Pollutants. Committee on the Medical Effects of Air Pollutants (COMEAP); 2010.

[39] Franco M, Ordunez P, Caballero B, Granados JAT, Lazo M, Bernal JL, et al. Impact of Energy Intake, Physical Activity, and Population-wide Weight Loss on Cardiovascular Disease and Diabetes Mortality in Cuba, 1980 2005. American Journal of Epidemiology. 2007 9;166:1374–80. Available from: https://academic.oup.com/aje/article-lookup/doi/10.1093/aje/kwm226.

[40] Ornish D. Intensive Lifestyle Changes for Reversal of Coronary Heart Disease. JAMA. 1998 12;280:2001. Available from: http://jama.jamanetwork.com/article.aspx?doi=10.1001/jama.280.23.2001.

[41] Capewell S, O’Flaherty M. Rapid mortality falls after risk-factor changes in populations. The Lancet. 2011 8;378:752–3. Available from: https://linkinghub.elsevier.com/retrieve/pii/S0140673610623021.

[42] Capewell S, O’Flaherty M. Can dietary changes rapidly decrease cardiovascular mortality rates? European Heart Journal. 2011 5;32:1187–9. Available from: https://academic.oup.com/eurheartj/article-lookup/doi/10.1093/eurheartj/ehr049.

[43] Wang Y, Li F, Wang Z, Qiu T, Shen Y, Wang M. Fruit and vegetable consumption and risk of lung cancer: A dose-response meta-analysis of prospective cohort studies. Lung Cancer. 2015 5;88:124–30.

[44] Harashima E, Nakagawa Y, Urata G, Tsuji K, Shirataka M, Matsumura Y. Time-lag estimate between dietary intake and breast cancer mortality in Japan. Asia Pacific Journal of Clinical Nutrition. 2007;16:193–8.

[45] Tsuji K, Harashima E, Nakagawa Y, Urata G, Shirataka M. Time-lag effect of dietary fiber and fat intake ratio on Japanese colon cancer mortality. Biomedical and environmental sciences : BES. 1996 9;9:223–8.

[46] Lin HH, Murray M, Cohen T, Colijn C, Ezzati M. Effects of smoking and solid-fuel use on COPD, lung cancer, and tuberculosis in China: a time-based, multiple risk factor, modelling study. The Lancet. 2008;372:1473–83. Available from: https://pubmed.ncbi.nlm.nih.gov/18835640/.

[47] Moutet L, Bigo A, Quirion P, Temime L, Jean K. Different pathways toward net-zero emissions imply diverging health impacts: a health impact assessment study for France. Environmental Research: Health. 2024;2. Available from: 10.1088/2752-5309/ad5750.

[48] Toujgani H, Berlivet J, Berthy F, Allès B, Brunin J, Fouillet H, et al. Dietary pattern trajectories in French adults of the NutriNet-Santé cohort over time (2014–2022): role of socio-economic factors. British Journal of Nutrition. 2024 11;132:1184–93. Available from: https://www.cambridge.org/core/product/identifier/S0007114524002514/type/journal_article.

[49] Kesse-Guyot E, Pointereau P, Brunin J, Perraud E, Toujgani H, Berthy F, et al. Trade-offs between blue water use and greenhouse gas emissions related to food systems: An optimization study for French adults. Sustainable Production and Consumption. 2023 11;42:33–43. Available from: https://linkinghub.elsevier.com/retrieve/pii/S2352550923002154.

[50] Gehring J, Touvier M, Baudry J, Julia C, Buscail C, Srour B, et al. Consumption of Ultra-Processed Foods by Pesco-Vegetarians, Vegetarians, and Vegans: Associations with Duration and Age at Diet Initiation. The Journal of Nutrition. 2021 1;151:120–31. Available from: https://linkinghub.elsevier.com/retrieve/pii/S0022316622000037.

[51] Kesse-Guyot E, Allès B, Brunin J, Langevin B, Fouillet H, Dussiot A, et al. Environmental pressures and pesticide exposure associated with an increase in the share of plant-based foods in the diet. Scientific Reports. 2023 11;13:19317. Available from: https://www.nature.com/articles/s41598-023-46032-z.

[52] Willett W, Rockström J, Loken B, Springmann M, Lang T, Vermeulen S, et al. Food in the Anthropocene: the EAT–Lancet Commission on healthy diets from sustainable food systems. The Lancet. 2019 2;393:447–92. Available from: https://linkinghub.elsevier.com/retrieve/pii/S0140673618317884.

[53] Laine JE, Huybrechts I, Gunter MJ, Ferrari P, Weiderpass E, Tsilidis K, et al. Co-benefits from sustainable dietary shifts for population and environmental health: an assessment from a large European cohort study. The Lancet Planetary Health. 2021 11;5:e786–96. Available from: https://linkinghub.elsevier.com/retrieve/pii/S2542519621002503.

